# Twelve Distinct Laboratory Methods Used to Measure SARS-CoV-2 in Wastewaters throughout a Three-Year Ontario-Wide, Canada Study: Impact on Public Health Interpretation of Disease Incidence

**DOI:** 10.64898/2026.03.27.26349524

**Authors:** Nada Hegazy, K. Ken Peng, Johanna de Haan-Ward, Elizabeth Renouf, Elizabeth Mercier, Shen Wan, X. Joan Hu, Charmaine Dean, Mark Servos, Elizabeth Edwards, Gustavo Ybazeta, Marc Habash, Lawrence Goodridge, R. Stephen Brown, Sarah Jane Payne, Andrea Kirkwood, Christopher Kyle, R. Michael McKay, Kimberly Gilbride, Christopher DeGroot, Robert Delatolla

## Abstract

Wastewater and environmental monitoring (WEM) was a critical public health surveillance tool for SARS-CoV-2 surveillance during the COVID-19 Pandemic. However, substantial methodological heterogeneity across laboratories continues to challenge the interpretation and thus compromise the actionability of resulting WEM measurements. This study quantifies interlaboratory concordance in SARS-CoV-2 WEM measurements using influent wastewater samples collected between September 2021 and January 2024 at a single wastewater treatment facility within the Ontario Wastewater Surveillance Initiative, analyzed independently by 12 laboratories using their routine methods. In the absence of a known true viral concentration, interlaboratory WEM measurements were evaluated against a facility-specific longitudinal benchmark derived from routine surveillance at the source facility and correlated to clinical surveillance metrics. Concordance was assessed across four WEM measurement units commonly used in practice: SARS-CoV-2 copies/mL, SARS-CoV-2 copies/copies of PMMoV, and their standardized counterpart wastewater viral activity level (WVAL) units of WVAL-standardized SARS-CoV-2 copies/mL and WVAL-standardized SARS-CoV-2 copies/copies of PMMoV. Measurements in each unit were analyzed using complementary analytical frameworks, including categorical concordance metrics, principal component analysis, and linear mixed-effects modelling. Across the study period, interlaboratory measurements consistently captured benchmark temporal dynamics, including major peaks and periods of low activity, but showed substantial variation in magnitude and public-health interpretation across laboratory methods. Concordance was strongest during epidemiological extremes and deteriorated during transitional periods, increasing the risk of misclassification with potentially implications for public health decision-making. To explore potential laboratory methodological drivers of agreement, associations between the benchmark concordance and the laboratory-specific concentration, extraction, and RT-qPCR analytical steps were assessed using Fisher’s exact tests, alongside extracted-mass threshold analyses. No single methodological factor showed a statistically significant association with benchmark concordance in this study; however, several parameters, including RNA template volume, total RT-qPCR reaction volume, and extracted mass of analyzed settled solids, may warrant further investigation in future studies.

## 1. Introduction

Wastewater and environmental monitoring (WEM) has become an essential public health tool around the world for providing population-wide, real-time monitoring of SARS-CoV-2, influenza, respiratory syncytial virus (RSV), and other emerging pathogens ^1–9^. WEM provides reliable population-wide surveillance and is a leading indicator when clinical testing capacity becomes limited, enabling health authorities to monitor infection trends, detect upcoming waves earlier than clinical metrics, and support policy decisions in real time ^10–13^. Even after the end of the COVID-19 public health emergency, key global public health agencies like the World Health Organization (WHO), Centers for Disease Control and Prevention (CDC), the U.S. Environmental Protection Agency (US EPA), the Public Health Agency of Canada (PHAC), the U.K. Health and Safety Agency (UKHSA), and the National Institute for Public Health and the Environment, Netherlands (RIVM) have recognized WEM’s continued relevance, as it fills gaps left by declining clinical testing and changing surveillance priorities post-pandemic ^14–20^. As WEM expands to target more pathogens and new strategies are developed to integrate it into routine public health surveillance, ensuring the reliability and comparability of analytical methods used across laboratories is becoming increasingly important. This is essential to maintain consistent data quality, reduce inter-site variability, enable comparison of disease burden across geospatial locations and support meaningful data integration and trend analysis ^21–23^.

In Ontario, Canada, the Wastewater Surveillance Initiative (WSI) was launched in January 2021 by the provincial Ministry of the Environment, Conservation and Parks (MECP). The program coordinated SARS-CoV-2 monitoring across 107 sites at the peak of the program, with data being generated by 13 laboratories located in 13 independent academic institutions in the Province of Ontario. Each of the 13 laboratories employed a distinct method to analyze SARS-CoV-2 in the program, with the laboratories forming the Ontario Wastewater Surveillance Consortium (OWSC) to share information and coordinate efforts ^24^. To support data consistency across the 13 laboratories, recurrent interlaboratory analyses were conducted throughout the program under the coordination of the Ontario Clean Water Agency (OCWA) from February 2021 through July 2024 ^25^. An Ontario-specific interlaboratory study using endogenous wastewater samples collected from September 2021 through July 2024, based on a subset of the interlaboratory dataset, demonstrated that methodological differences across laboratories could yield broadly comparable measurements, while also highlighting the need for further investigation of method-specific biases ^26^.

A current barrier to confidence in WEM and the actioning of WEM across various geospatial locations remains the substantial methodological heterogeneity used to measure health targets in wastewaters. Differences in wastewater sample processing, solids concentration, nucleic acid extraction, normalization strategies, and RT-qPCR workflows have been cited as sources of bias that complicate disease interpretation ^6,13,27–30^. Inaccurate and inconsistent measurements in WEM can potentially yield contradictory public health messages, obscure true disease burden, and erode trust in the surveillance system ^31^. As WEM transitions to a long-term national and international surveillance strategy, these concerns underscore the need to not only understand between-laboratory differences introduced by the use of diverse analytical methods but also comprehend how such differences influence differences in public health interpretation.

As WEM has scaled from single-community surveillance to coordinated regional and national networks, interlaboratory comparability has emerged as a prerequisite for reliable surveillance ^32–34^. Prior interlaboratory studies, including studies that evaluated 36 methods from across the U.S. ^32^, eight Canadian laboratories ^35^, eight U.S. laboratories ^36^, the 13 Canadian laboratories participating in the Ontario WSI program with the Public Health Agency of Canada – National Microbiology Laboratory ^26^, and four laboratories in Italy ^37^ have demonstrated that, while diverse laboratory analytical methods and workflows can achieve acceptable within-lab reproducibility, between-method viral measurements for identical samples can differ by several orders of magnitude, resulting in divergent measurements across geospatial locations within the same coordinated surveillance frameworks. These discrepancies in measurements and, hence, subsequent interpretation of disease burden are currently inferred to be attributed to differences in sample concentration techniques, extraction protocols, and RT-qPCR procedures ^32,38^. Accordingly, organizations like the CDC’s National Wastewater Surveillance System (NWSS) and the WHO have emphasized that methodological diversity and limited standardization contribute to reduced data comparability and constrain the value of WEM for coordinated public health decision-making ^39,40^. Furthermore, most interlaboratory evaluations to date have used samples with spiked surrogate viruses, which directly alter the physicochemical and biological complexity inherent to targeting endogenous health targets in wastewater matrices. Consequently, spiked sample comparative studies provide limited insight into how the use of different laboratory methods affects the interpretation of WEM measurements for public health decision-making.

To address these gaps, this study aims to evaluate how differing routine laboratory methods influence public health interpretation of SARS-CoV-2 wastewater measurements based on an interlaboratory study of 13 endogenous samples collected from a single wastewater treatment facility and then analyzed by 12 distinct laboratory methods employed within the Ontario WSI program ^24^. Out of the 13 laboratories participating in the program, 12 laboratories have selected to opt in to this analysis. This study aims to examine whether laboratory methods classify the same samples into the same wastewater viral activity level, and whether deviations align with or diverge from a reference benchmark derived from the facility’s longitudinal wastewater record, which could enable the direct assessment of interpretation bias and provide insight into how method-driven differences shape public health-relevant outcomes. The reference benchmark used in this study was verified against the clinical case data set from the sewershed area where the wastewater was collected, demonstrating a 0.74 Spearman’s rank correlation to clinical cases during prominent Omicron waves. Between-method differences are further assessed using the complementary analytical frameworks, principal component analysis (PCA), and mixed-effects modelling, which together characterize concordance, multivariate pattern similarity, and variance components. This study introduces a structured analytical framework for comparing interlaboratory wastewater surveillance results of endogenous targets in the absence of a known true concentration. By integrating categorical concordance measures, PCA analyses, and mixed-effects modelling relative to a longitudinal benchmark that was validated against clinical data, the framework enables reproducible comparison of viral measurement interpretations across laboratory methods and locations, extending WEM from site-specific trend analysis toward cross-jurisdictional public health application. Accordingly, this manuscript aims to (i) assess whether laboratory methods influence the public health interpretation of WEM viral measurements represented as SARS-CoV-2 copies/mL, SARS-CoV-2 copies/copies of PMMoV, WVAL-standardized SARS-CoV-2 copies/mL, and WVAL-standardized SARS-CoV-2 copies/copies of PMMoV, and (ii) identify the analytical factors that most strongly contribute to between method-differences in these interpretations. By elucidating which components of the analytical process drive divergence in SARS-CoV-2 measurements between laboratory methods, this work supports the development of standardized methodological guidance for WEM programs and informs policy discussions related to the standardization of WEM practices for future pandemics and emerging pathogen surveillance.

## 2 Methodology

### 2.1 Overview of Ontario’s Interlaboratory Study Procedure

The Ministry of Environment, Conservation, and Parks (MECP), along with the Public Health Agency of Canada – National Microbial Laboratory (PHAC-NML) and the Ontario Clean Water Agency (OCWA), coordinated an interlaboratory initiative that collected 18 homogenized wastewater samples containing endogenous SARS-CoV-2 between February 1, 2021 and April 22, 2024 from an anonymized wastewater treatment plant in Ontario (hereafter referred to as the “source facility”). The source facility was known to be impacted by COVID-19 and was routinely monitored under the Ontario WSI program from January 1^st^, 2021, until July 31^st^, 2024 ^24^. The region contributing to the wastewater treated at the source facility was also routinely monitored for clinical cases of COVID-19 by a local Ontario public health unit and Public Health Ontario. Throughout the program, a total of 18 homogenized influent wastewater samples were collected and analyzed by participating laboratories across the province between February 1, 2021, and April 22, 2024. For this study, 13 samples collected between September 27, 2021, and January 29, 2024, were retained, excluding three early-program samples for which methodological inconsistencies in the Ontario WSI workflows were documented, and two later samples that were collected from a different source facility. These 13 retained samples are denoted by letters A through M for reference in subsequent analyses.

OCWA collected 500 mL grab influent wastewater samples and shipped a minimum of three biological replicates to all 12 participating laboratories on ice via overnight courier in insulated coolers ^25^. Upon receipt, participating laboratories stored samples at 4°C and conducted sample concentration and quantification of SARS-CoV-2 N1 and N2 gene regions and pepper mild mottle virus (PMMoV) within 48 hours of collection by means of RT-qPCR ^24,25^. Participating laboratories analyzed these aliquots using the distinct in-house analytical methods that they applied for routine SARS-CoV-2 surveillance throughout the three-year span of the program, including their established procedures for enrichment and concentration, nucleic acid extraction, RT-qPCR amplification, and normalization. These 12 methods used for concentration, nucleic acid extraction, and RT-qPCR amplification are described in detail in D’Aoust el al. (2024) ^24^. Summaries of the methods are also presented in Tables S1 and S2 of the Supplementary Material. All laboratories homogenized the interlaboratory samples prior to processing, analyzed a minimum of three biological replicates for each interlaboratory collection date, and analyzed a minimum of three technical replicates for each biological replicate of each SARS-CoV-2 gene target, N1 and N2. The participating laboratories reported the final SARS-CoV-2 concentrations (a minimum of 9 measurements, 3 biological replicates by 3 technical triplicates) as N1-N2 copies/mL of wastewater, where N1-N2 was the average of the N1 and N2 measurements, with PMMoV concentrations also provided and used for normalization. No proficiency or reproducibility test was assigned, as the interlaboratory study design intentionally focuses on interpretability under realistic endogenous matrix conditions.

#### 2.1.1 Data Collection and Processing

For all 13 interlaboratory samples included in this study, participating laboratories reported raw SARS-CoV-2 N1 and N2 gene target measurements for each gene target in copies/mL. Across all samples, laboratories generated between 3 and 7 biological replicates (mean 3.65 ± 1.26 biological replicates), and including triple technical replicate data, produced 1692 observations for the N1 gene target and 1620 observations for the N2 gene target. The difference in observation counts of the N1 and N2 gene targets is attributed to the fact that not all laboratories measured both N1 and N2 gene targets for every interlaboratory sample, and that some laboratories preferentially quantified the N1 gene target alongside alternative targets such as N200 or the E gene target ^41^. When a technical replicate yielded no detectable measurement for a given target, the laboratory-specific assay limit of detection (ALOC) was substituted; this adjustment was applied to 54 observations. To obtain the quantitative viral measurement estimates, technical replicates and respective N1 and N2 gene target measurements within each well were averaged, producing three per-well means for each biological replicate as N1-N2 copies/mL. In cases where only one SARS-CoV-2 target gene (N1 or N2) was quantified for a given sample due to various reasons applicable to analyzing samples during a pandemic, that single target concentration was used as the estimated N1-N2 copies/mL; this was applied to 180 observations ^42^. Biomarker-normalized concentrations (PMMoV-normalized N1-N2, hereafter referred to as N1-N2 copies/copies of PMMoV) were calculated by dividing each N1-N2 copies/mL estimate by the mean PMMoV concentration for the corresponding biological replicate.

In addition to the raw and biomarker-normalized concentrations, this study further incorporates the Wastewater Viral Activity Level (WVAL) to evaluate a standardized, public health-oriented metric that is actively applied in WEM programs in the U.S., as per the Centre for Disease Control and Prevention ^43^ and Delatolla et al. (2025) ^44^. WVAL is a unitless index developed by the U.S. CDC to standardize SARS-CoV-2 viral measurement across various locations, enabling the data to become geospatially comparable ^43,44^. However, to date, the extent to which WVAL harmonizes results across laboratories employing different concentration, extraction, and amplification methods has not been fully characterized. As such, incorporating WVAL into this study permits the evaluation of its effect on comparability among laboratories or whether laboratory-specific methodological differences persist despite standardization. For each participating laboratory, longitudinal benchmark data, and WEM historical data (explained further in Section 2.3.1), the WVAL was computed from the baseline (10th percentile) and standard deviation of the log-transformed WEM viral measurement and is updated every six months to include new data. Although WVALs are routinely updated every six months under surveillance conditions, the non-longitudinal nature of the interlaboratory samples in this study required the computation of a single baseline and standard deviation for the entire study period. In this study, WVALs were calculated for both non-normalized concentrations (WVAL-standardized - N1-N2 copies/mL) and biomarker-normalized concentrations (WVAL standardized - N1-N2 copies/copies of PMMoV).

The benchmark WEM data from the source facility were treated analogously. Technical triplicates were available for SARS-CoV-2 N1, N2, and N200 gene targets as well as from PMMoV normalization biomarker, where N200 gene targeting was used by a single laboratory to replace the N1 gene target ^41^. For each triplicate, similarly to the interlaboratory data, the mean of the target genes was determined using the available targets for the sampling date (i.e., mean of N1 and N2; mean of N2 and N200; or a single N1 or N2 target when only one was quantified). Note that throughout this manuscript, the mean of these gene targets is referred to as N1-N2 copies/mL, regardless of the target genes included in the calculation. As for the biomarker-normalized benchmark data, they were computed by dividing these means by the corresponding PMMoV mean for each corresponding sample date. The WVAL-standardized measurements (in both N1-N2 copies/mL and N1-N2 copies/copies of PMMoV) were computed for the benchmark data. The baseline 10th percentile and standard deviation were computed using all observations collected after the first interlaboratory sample considered for this study was collected (September 27, 2021). Throughout this study, the interlaboratory samples were compared against the 7-day midpoint average measurements from the benchmark, as the benchmark in this study serves to represent a locally consistent reference signal rather than a true concentration.

After combining the benchmark and the interlaboratory data, the final dataset used in this study consisted of 1789 observations of N1-N2 copies/mL, and 1774 observations PMMoV-normalized observations of N1-N2 copies/copies of PMMoV, with differences in datasets attributed to one participating laboratory that did not report PMMoV genomic copy measurements for 4 biological replicates of one interlaboratory sample (Sample A), and one biological replicate of another sample (Sample B), which precluded calculation of the normalized metric for those observations. Accordingly, analyses in this manuscript were conducted using (i) non-normalized SARS-CoV-2 N1-N2 concentration per mL of sample (N1-N2 copies/mL of sample volume), ii) PMMoV normalized concentrations (N1-N2 copies/copies of PMMoV), (iii) WVAL-standardized N1-N2 copies/mL and (iv) WVAL-standardized N1-N2 copies/copies of PMMoV.

### 2.2 Longitudinal Benchmark for Interlaboratory Comparison

In this study, the endogenous wastewater samples sent to the participating Ontario WSI laboratories lack traceable “true” SARS-CoV-2 N1 and N2 viral measurements, as one of the primary purposes of the inter-laboratory analysis was to allow the participating laboratories to demonstrate their independent ability to produce credible results. Due to a lack of proficiency testing in the interlaboratory analysis and the lack of a “true” concentration for endogenous wastewater samples throughout the interlaboratory analysis, a reference benchmark was defined using a longitudinal wastewater dataset derived from the same source facility that generated all interlaboratory samples included in this study. The source facility conducted routine SARS-CoV-2 wastewater surveillance as part of the Ontario WSI program, with N1-N2 measurements collected on average three times per week across the study period ^24^. Although this benchmark does not represent a true concentration, it provides an internally consistent, facility-specific reference that reflects the expected temporal pattern of SARS-CoV-2 viral activity at each interlaboratory sampling point. The benchmark wastewater measurement for this study was validated against clinical case data provided by the Ontario Ministry of Health (MOH) from the same sewershed, yielding a strong Spearman’s correlation of 0.74 during the Omicron BA.1 and BA.2 waves period prior to the decline in population-wide PCR testing ^24^. Accordingly, the benchmark serves as a practical reference against which interlaboratory interpretations can be assessed, providing the common frame required to compare the SARS-CoV-2 N1-N2 copies/mL, N1-N2 copies/copies of PMMoV, and WVAL-standardized units across laboratories throughout this study.

### 2.3 Statistical Methodology

To evaluate the proportion of deviation that laboratory-specific methodological choices drive the interpretation and magnitude of SARS-CoV-2 WEM measurements, this study applies a multi-layered analytical framework combining categorical concordance assessment, multivariate pattern analysis, and hierarchical statistical modelling. First, the wastewater viral activity level index categorization was performed to translate non-normalized and normalized SARS-CoV-2 measurements into public-health-relevant activity categories, enabling direct comparison between each laboratory and the benchmark. This categorical analysis identifies where interpretation diverges, which inter-methodological samples drive potential disagreement, and which methods remain aligned or deviate under real-world endogenous conditions. Second, PCA was conducted to provide further assessment of how far methods deviate from the benchmark in the reduced-dimension space. Third, mixed-effects modelling was performed to evaluate whether method identity significantly influences measured target concentrations while accounting for nested study design (biological replicates within samples and methods). Together, these analyses aim to capture interpretive disagreement, structural measurement variability, and statistically significant effects caused by the 12 participating laboratories and their distinct methods to provide a comprehensive assessment of method-driven differences across the interlaboratory dataset.

#### 2.3.1 Wastewater Activity Level Index – Quantile-Based Classification

Many public health agencies around the world summarize WEM measurements into discrete “activity levels” (e.g., low, moderate, high, very high) using percentile or quantile-based thresholds to assess viral measurement trends, and to support dashboard communication and decision-making. Such categorical approaches have been used in Canada’s national WEM dashboards and in U.S. programs to contextualize local and national trends, allocate resources, and provide consistent public-facing risk categories while mitigating measurement error and uncertainty ^45–48^.

In this study, to compare interlaboratory measurements using public-health-relevant measurement categories, SARS-CoV-2 N1-N2 measurements were mapped to four wastewater viral activity levels: “low”, “moderate”, “high”, and “very high” using a quartile-based classification framework. This quartile-based approach was applied separately to (i) the benchmark reference longitudinal data and (ii) historical Ontario WSI 7-day midpoint average datasets used to contextualize each participating laboratory relative to their respective routine WSI sites that were under the provincial surveillance program ^24^. For the benchmark dataset, to avoid undue influence of extreme values on percentile boundaries, outliers were identified using the Interquartile Range (IQR) method from the daily N1-N2 copies/mL concentrations and removed, resulting in the identification and exclusion of 53 outliers out of n = 717 longitudinal data points. Quartiles (25^th^, 50^th^, and 75^th^ percentiles) were then calculated from the 7-day midpoint average N1-N2 copies/mL and N1-N2 copies/copies of PMMoV. These facility-specific quartiles defined the reference activity categories against which interlaboratory classifications of the shared endogenous samples were evaluated.

In parallel, quartiles were computed from historical WSI 7-day midpoint average data for each laboratory’s routine sampling locations that participated under the Ontario WSI program from January 1^st^, 2021, until the decommissioning of the program on July 30^th^, 2024, with an average of 3 samples per week ^24^. These laboratory-specific historical quartiles were used to assign wastewater viral activity levels in a manner consistent with how each participating laboratory’s WSI data are interpreted operationally. In some cases, because the WSI sites differ in sewershed population, flow regime, and disease burden from the source-facility where the interlaboratory samples were collected, three out of the 12 corresponding routine sampling sites (corresponding to laboratories σ, τ, and υ) exhibited historical measurement distribution that did not align with the magnitude range of the interlaboratory samples. In such instances, the historical WSI distributions in N1-N2 copies/mL (corresponding to laboratories σ and τ) were scaled by 5.6 and 11.7, respectively, and the N1-N2 copies/copies of PMMoV distribution was scaled by 28 (corresponding to laboratory υ) in order to align with the magnitude range of the interlaboratory samples prior to quartile calculation to avoid spuriously inflated or deflated activity classifications driven solely by site-specific measurement magnitude.

For both the benchmark dataset and each laboratory’s historical WSI dataset, measurements were categorized using the following scheme: low measurement < 25^th^ percentile, moderate measurement between 25^th^ and 50^th^ percentile, high measurement between 50^th^ and 75^th^ percentile, very high ≥ 75^th^ percentile. This four-tier structure provides finer resolution than commonly used three-level national frameworks, and is a similar framework to those applied in public health units in Ontario ^46,49^. While existing WEM programs actively utilize percentile-based or quantile-based categorization of wastewater measurements for dashboards and operational decision support, to date, interlaboratory studies have largely focused on concentration and comparability rather than explicit concordance in categorical activity level assignments.

##### 2.3.1.1 Cohen’s Kappa (к)

In order to quantify weighted agreement of the wastewater viral activity level index for each laboratory and the benchmark, quadratic weighted Cohen’s Kappa (к) was calculated, which accounts for the ordinal structure of the wastewater viral activity categorization, and penalizes larger category discrepancies more heavily than minor classification disagreements. Values of к were interpreted as follows: к < 0.00 indicating poor agreement, 0.00 – 0.20 indicating slight agreement, 0.21 – 0.40 indicating fair agreement, 0.41 – 0.60 indicating moderate agreement, 0.61 – 0.80 indicating substantial agreement, and 0.81 – 1.00 indicating almost perfect agreement ^50^.

#### 2.3.2 Principal Component Analysis (PCA)

Building on the descriptive analyses, PCA was applied to compare participating laboratories against the benchmark dataset. For each laboratory and sample, the biological replicates were averaged to generate laboratory-level summaries for the four viral measurement units (N1-N2 copies/mL, N1-N2 copies/copies, WVAL-standardized N1-N2 copies/mL, and WVAL-standardized copies/copies). To ensure that comparisons reflected cross-sample variation patterns rather than absolute concentration magnitudes, all non-WVAL-standardized measurements (N1-N2 copies/mL and N1-N2 copies/copies of PMMoV) were centred and scaled to zero mean and unit variance prior to conducting the PCA. PCA then transformed the original lab measurements into orthogonal (uncorrelated) components that capture the largest proportions of variation among the data, highlighting the samples most strongly driving differences across laboratories. The analysis was done using the R package FactoMineR ^51^, with R version 4.4.3 ^51,52^.

To quantify each laboratory’s deviations from the benchmark, the Euclidean distance between laboratory scores and benchmark scores using the first three principal components (PCs) was calculated, and laboratories were ranked from closest to farthest. The choice of the first three PCs was made a priori to balance parsimony with coverage of variability. This approach yields a single, multivariate measure of similarity that accounts for correlations across the interlaboratory samples. The resulting Euclidean distance with the first 3 PCs condenses complex multi-lab data into one interpretable metric while reducing noise through dimension reduction. However, results are sensitive to preprocessing choices and the number of PCs retained. PCA primarily captures linear patterns, and averaging biological replicates removes within-laboratory variability. These limitations motivate the subsequent mixed-effects modelling, which explicitly incorporates replicate-level structure and provides a complementary assessment of laboratory-driven deviations.

#### 2.3.3 Regression with Mixed Effects Model

A linear mixed effects model was fitted to quantify the extent to which the laboratory performing the analysis influenced measured viral measurement and their corresponding WVAL-standardized units, while accounting for the study design. The response variable was the viral measurement represented by the four units discussed above. Because the SARS-CoV-2 WEM data are right-skewed, the interlaboratory measurements and the benchmark were log-transformed prior to modelling. The fixed effects component consists of the 12 participating laboratories in the interlaboratory study and the benchmark (a total of 13 levels), resulting in 13 estimated coefficients relative to the benchmark, one for each laboratory, with the intercept representing the benchmark. A nested structure is used for the random effects, such that the technical triplicates were nested within biological replicates, which were in turn nested within each interlaboratory sample.

Four separate models, one for each viral measurement metric, similarly to the PCA (N1-N2 copies/mL, N1-N2 copies/copies of PMMoV, WVAL-standardized N1-N2 copies/mL, and WVAL-standardized N1-N2 copies/copies of PMMoV), were fitted using the R package lme4, version 1.1.37 ^53^ with R version 4.4.3 ^52^. An ANOVA F-test was conducted for each model using the R package nlme, version 3.1.167 ^54^, to assess the null hypothesis that the laboratory, the sole fixed effect in this model, does not significantly improve the fit of the data to the model, compared to that of the intercept-only model. For the mixed-effects model component, benchmark daily measurements were matched to interlaboratory dates using the same-day daily measurement, or, when unavailable, the nearest adjacent date (± 1 day).

#### 2.3.4 Association Between Laboratory Methodological Drivers and the Concordance with the Benchmark

To explore whether inter-laboratory methodological differences contributed to variation in concordance with the benchmark, a broad laboratory-level screening analysis was conducted in this study. Each laboratory was classified as having acceptable concordance (good or moderate) or poor concordance with the benchmark based on its overall pattern across all sample-level analyses. Associations between laboratory performance category and the following methodological characteristics were then evaluated using Fisher’s exact tests ^55^, which are appropriate for assessing associations between categorical variables in small-sample settings. The following methodological variables were assessed: concentration method (filtration or centrifugation), centrifugation speed (if applicable) for both spins, standardized materials in assays for the N1 and N2 gene regions, N1 and N2 RNA template volume (μL), N1 and N2 total RT-qPCR reaction volume (μL), N1 and N2 RT-step at 25°C, cDNA synthesis duration (min), initial denaturation duration (min), cycling number, cycling denaturation duration (s), and cycling annealing extension (°C), using the Fisher’s exact test (Tables S1 and S2 in Supplementary Material). For each methodological variable, the test was conducted only when the corresponding method was consistent within laboratories over the study period; variables for which any laboratory reported more than one value over time were excluded from testing. For numeric variables, values were summarized at the laboratory level and dichotomized as above or below the overall laboratory mean before the analysis. This screening analysis provides a first assessment of potential methodological drivers, noting that the available metadata were only summarized at the laboratory level rather than at the sample level.

Given these limitations, a more targeted assessment was performed, focusing on extracted mass/pellet weight, used for the concentration stage of the SARS-CoV-2 N1, N2, and PMMoV quantification, as it was the sole methodological variable at the sample level. Although laboratories did not record direct pellet weights, 11 out of the 12 laboratories that used centrifugation or PEG for their respective solids concentration method have reported centrifuged sample volume; extracted mass was thus estimated using the recorded total solids (TS) (mg/L) measured from the source facility on the corresponding interlaboratory sample days, multiplied by the centrifuged volume (L). The estimated extracted mass ranged from 28.64 to 250.0 mg (94.64 ± 60.91 mg; n=11) (Table S1 in Supplementary Material).

Each laboratory-sample combination was classified as either a match or a non-match to the benchmark based on the quantile-based measurement categorization described in Section 2.3.1. Extracted mass was then dichotomized into “low” (≤ K) or “high” (> K) across a sequence of thresholds K ranging from 20 to 200 mg. For each value of K, a chi-square test of independence^56^ was conducted to evaluate whether the extracted mass category was associated with the benchmark match status. Given the sufficient number of laboratory-sample combinations, the chi-square approximation was used, and the results were consistent with those obtained using Fisher’s exact test. To summarize the sensitivity of this association to the chosen threshold K, p-values were plotted as a function of K, generating curves for each of the two viral measurement metric units explored (N1-N2 copies/mL, N1-N2 copies/copies PMMoV) based on quantile-based measurement categorization shown in Figure 2. This approach allows examination of whether extracted mass exhibits a stable, biologically meaningful relationship with benchmark concordance across a plausible range of cutoff definitions.

## 3 Results and Discussion

### 3.1 Interlaboratory Measurements and Benchmark

Throughout the study period (September 9^th^, 2021 – January 29, 2024), the benchmark 7-day midpoint average N1-N2 copies/mL viral measurements ranged between 0.96 to 664 N1-N2 copies/mL (mean = 118 ± 111 N1-N2 copies/mL; n = 855) and 9.3 x 10^-6^ and 6.4 x 10^-3^ 7-day midpoint average N1-N2 copies/copies of PMMoV (mean = 1.4 x 10^-3^ ± 1.2 x 10^-3^ N1-N2 copies/copies of PMMoV; n = 855) (Figure 1). The consecutive 7-day midpoint average WVAL-standardized N1-N2 copies/mL and WVAL-standardized N1-N2 copies/copies of PMMoV ranged between 0.02 and 25.5 (mean = 5.7 ± 5.6; n = 855) and 0.01 and 27.5 (mean = 6.0 ± 5.7; n = 855), respectively (Figure S1 in Supplementary Material). The WVAL-standardized measurement exhibited more consistent ranges, with measurements ranging between 0.02 – 25.5 for the WVAL-standardized N1-N2 copies/mL and 0.01 – 27.5 for the WVAL-standardized N1-N2 copies/copies of PMMoV, compared to the corresponding unstandardized ranges of 0.96 – 664 N1-N2 copies/mL and 9.3 x 10^-6^ – 6.4 x 10^-3^ N1-N2 copies/copies of PMMoV, while retaining their respective temporal dynamic, which is expected due to the nature of the WVAL-standardization procedure^43,44^.

**Figure 1:**
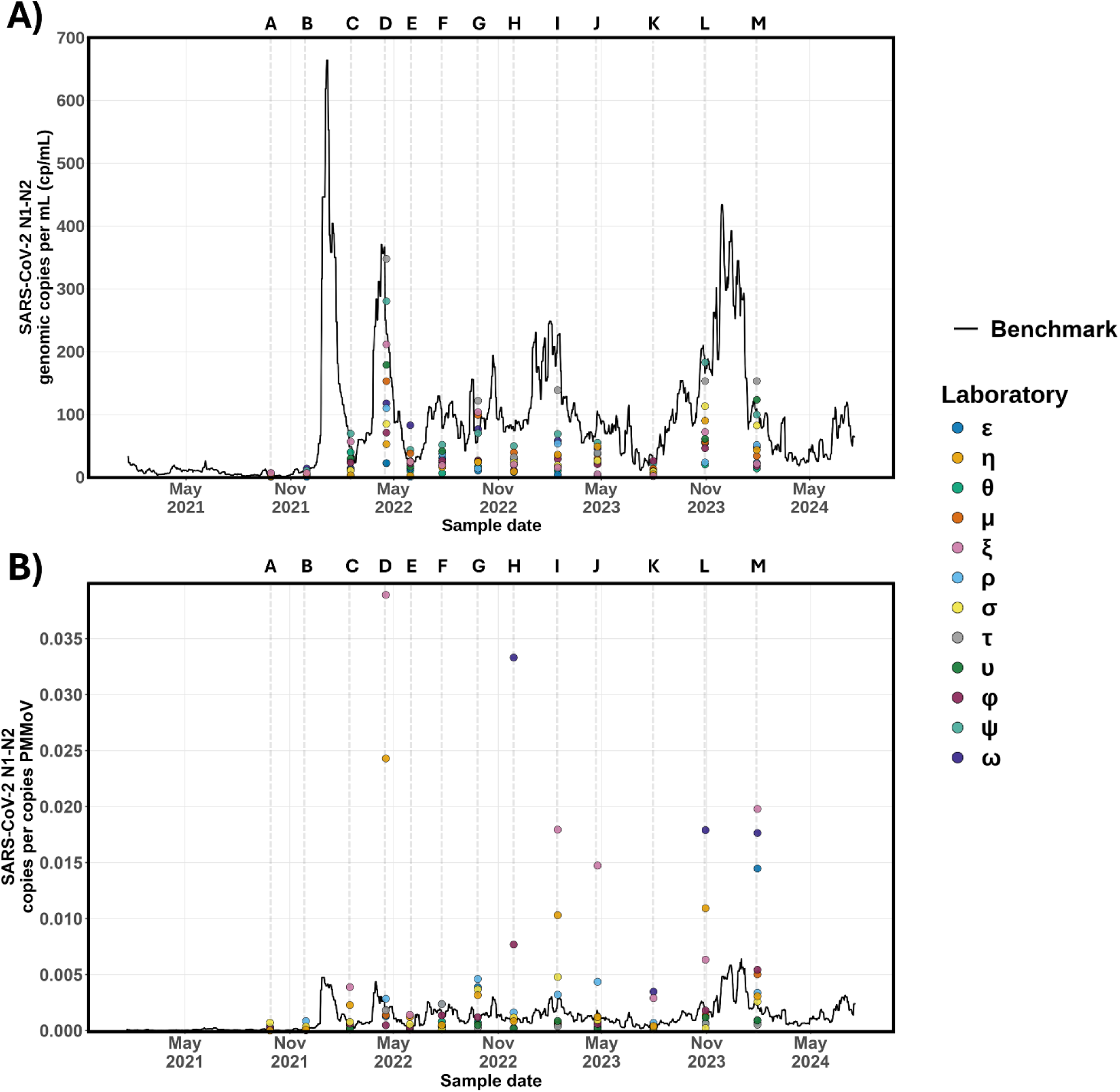
Inter-laboratory viral measurement relative to the benchmark measurement from the source facility. The solid black line represents the baseline measurement, defined as the 7-day moving average of A) N1-N2 copies/mL and B) N1-N2 copies/copies of PMMoV from the benchmark source facility from which the inter-laboratory samples were collected. Colored circles represent measurements reported by the 12 participating laboratories analyzing wastewater samples collected from the same location. Sample letters above the plotting area correspond to individual inter-laboratory sampling rounds. It is noted that some laboratories used different PMMoV standards compared to the source facility, hence creating a large difference between some laboratory measurements and the source facility measurements.

Across all four measurement units, the interlaboratory measurements from the 12 participating laboratories effectively capture the temporal dynamics observed in the benchmark data, including major epidemic peaks and periods of low incidence (Figure 1 and Figure S1 in Supplementary Material). Despite this agreement in trend detection, substantial divergence in absolute measurement magnitude was observed. For N1-N2 copies/mL, interlaboratory measurements spanned approximately two orders of magnitude across the study period, with laboratory-specific minima ranging between 0.74 and 5.7 N1-N2 copies/mL (a factor of 7.7) and maxima ranging between 40.0 to 348 N1-N2 copies/mL (a factor of 8.7), even though all measurements were derived from the same source facility during sampling rounds A through M (Figure 1A). Previous nationwide interlaboratory studies have demonstrated that different participating laboratories, despite each yielding reproducible results with their respective analytical methods for extraction, concentration, and RT-qPCR of SARS-CoV-2 viral particles, exhibited measurements that differ from each other by several orders of magnitude ^32,57^.

For N1-N2 copies/copies of PMMoV, the divergence in magnitude relative to the benchmark was attenuated but persisted, with laboratory-specific minima ranging between 2.4 x 10^-5^ and 6.8 x 10^-4^ N1-N2 copies/copies of PMMoV (a factor of 28.3), and maxima ranging between 1.4 x 10^-3^ and 6.7 x 10^-2^ N1-N2 copies/copies of PMMoV (a factor of 48.9) (Figure 1B). However, it is visually illustrated that the PMMoV-normalized, N1-N2 copies/copies of PMMoV are not visually aligned with the benchmark measurements to the same extent as the raw N1-N2 copies/mL (Figure 1). A likely explanation is that each participating laboratory used their own in-house standard material for PMMoV quantification, producing substantial divergence in reported PMMoV magnitudes from all participating laboratories, ranging from 133 up to 2.8 x 10^5^ PMMoV copies (a factor of 2100) (Figure S2 in Supplementary Material). This divergence in magnitude for PMMoV copies was documented previously and was attributed to heterogeneous standard materials and calibration practices ^36,58,59^.

### 3.2 Deviation between Viral Measurement Activity Classification and Benchmark

Classification of endogenous SARS-CoV-2 measurements into viral activity levels of low, moderate, high and very high revealed agreement levels between the laboratory methods that ranged from none to slight agreement (к = 0.23 in N1-N2 copies/mL and к = 0.13 N1-N2 copies/copies of PMMoV) to almost perfect agreement (к = 0.80 in N1-N2 copies/mL and к = 1.00 in N1-N2 copies/copies of PMMoV) (Figure 2). Samples A and D demonstrated the highest agreement across all participating laboratories, with agreements of к = 0.92 and к = 0.83 for N1-N2 copies/mL for each sample, respectively (Figure 2A), and к = 1.00 for both samples in N1-N2 copies/copies of PMMoV (Figure 2B). Sample A corresponds to a low-viral measurement period, where nearly all laboratories assigned a “low” viral measurement activity category. Sample D, the highest viral measurement across the entire interlaboratory period (Figure 1), was consistently classified as “very high” by 10 out of the 12 in N1-N2 copies/mL (Figure 2A) and 12 out of 12 in N1-N2 copies/copies of PMMoV (Figure 2B). These extremes in viral measurement activity levels reduced the influence of methodological impacts on viral measurement activity categorization.

**Figure 2:**
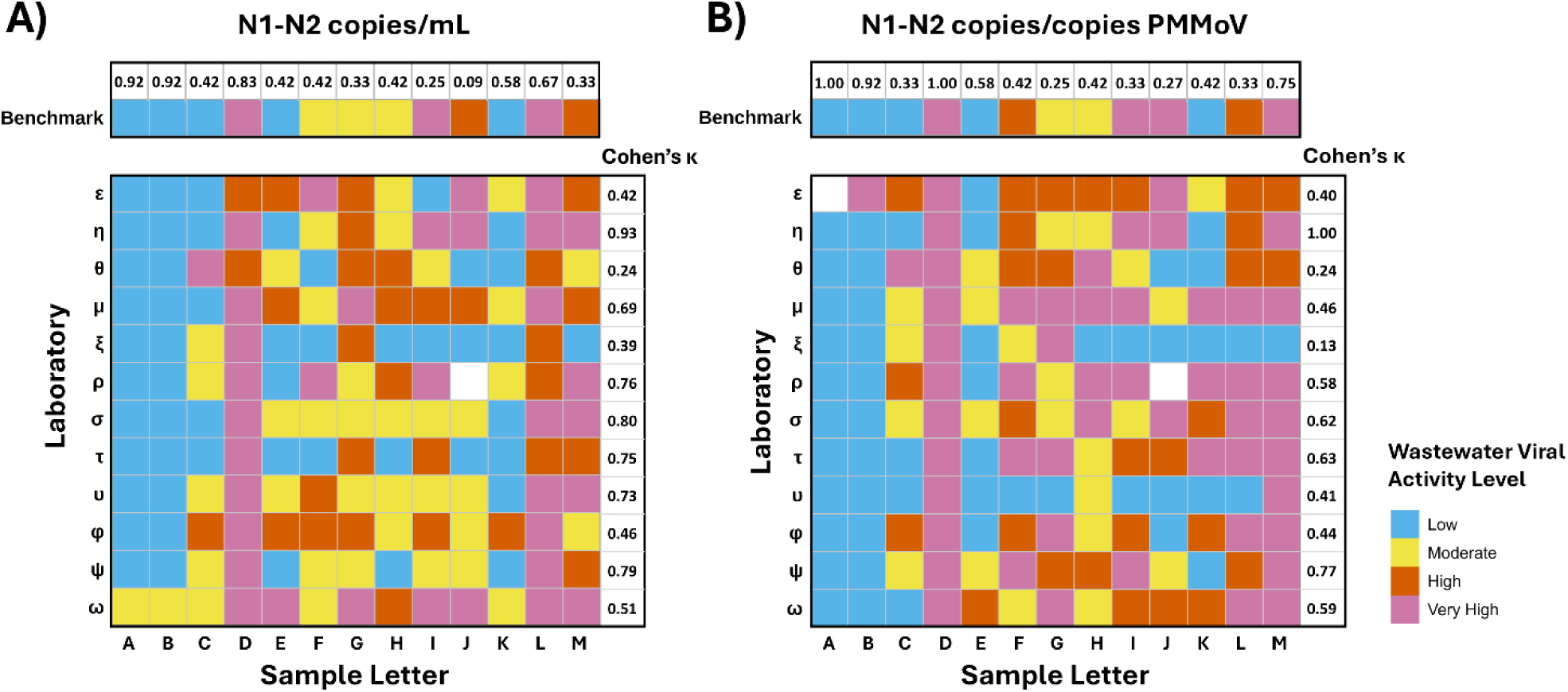
Wastewater viral activity level classifications for 13 endogenous wastewater samples compared across participating 12 laboratories and the benchmark reference series for A) non-normalized N1-N2 copies/mL and B) PMMoV-normalized N1-N2 concentration (N1-N2 copies/copies of PMMoV). Each tile represents the categorical viral activity (low, moderate, high, or very high) assigned by a laboratory for a given sample, with benchmark categories shown on the top row of the panel. On the right column of each panel are the Cohen’s к agreement scores to quantify the proportion of exact weighted matches of each laboratory with the benchmark. On the top row of each panel is the agreement ratio to quantify the proportion of exact matches of each interlaboratory sample.

In contrast, transitional periods between April 14, 2022, until January 29, 2024, yielded mixed classifications, as benchmark measurements oscillated from low to very high (Figure 1). During these periods, participating laboratories similarly reported mixed classifications (low, moderate, high, or very high) for the same interlaboratory sample. In this period, despite this variability, a subset of laboratories consistently aligned with the benchmark measurement. Specifically, laboratories η, μ, σ ρ, τ, υ, ψ and ω exhibited moderate to almost perfect agreements (к = 0.688 – 0.929) in N1-N2 copies/mL (Figure 2A), and laboratories η, ρ, τ, ψ, σ, and, ω exhibiting moderate to almost perfect agreements (к = 0.578 – 1.00) in N1-N2 copies/copies of PMMoV (Figure 2B). On the other hand, laboratories ε, θ, ξ, and φ exhibited only slight to fair agreement (к = 0.239 – 0.457) in N1-N2 copies/mL (Figure 2A), and laboratories ε, θ, μ, ξ, υ, and φ exhibited no agreement to fair agreement (к = 0.133 – 0.459) in N1-N2 copies/copies of PMMoV (Figure 2B). This transitional period corresponded to Ontario’s Omicron-driven hospitalization surges from 2022 to 2024 ^60,61^. Systemic bias and imprecision in quantified viral measurements can result in misclassification of activity levels, leading to flawed public health messaging, delayed alerts from false negatives or unnecessary resource allocation from false positives, thereby undermining public health decision-making ^62,63^. These disagreements which are likely driven by method-specific processing, extraction, and quantification differences, underscore the need to identify key methodological drivers of these disagreements. To further contextualize the strength of agreement with the benchmark in terms of overall measurement structure rather than categorical classification, PCA analyses were next applied to assess the differences between laboratory methods’ respective viral measurements relative to the benchmark.

In this study, this analysis was not applied to the WVAL-standardized N1-N2 copies/mL and WVAL-standardized copies/copies, as the quartile-based classification framework to the non-standardized N1-N2 copies/mL and N1-N2 copies/copies of PMMoV encodes a similar purpose as to the WVAL-standardization of representing the relative viral activity level ^43^.

### 3.3 PCA-based Divergence of Laboratories from the Benchmark

This study further quantifies the multivariate differences between the participating laboratory methods and the benchmark measurement in all four measurement units (N1-N2 copies/mL, N1-N2 copies/copies of PMMoV, WVAL-standardized N1-N2 copies/mL, and WVAL-standardized N1-N2 copies/copies of PMMoV). A clear pattern appearing from these plots is that the relative positioning of laboratories in the PCA space is highly sensitive to the choice of measurement unit (Figure 3 and Figure S3 in Supplementary Material). Both the overall spread of laboratory methods and their distances to the benchmark vary substantially across the four units, indicating that different units emphasize different aspects of multivariate variation. The laboratories highlighted as among the five closest to the benchmark (red) also differ notably across the four units used for comparison in this study: N1-N2 copies/mL, N1-N2 copies/copies of PMMoV, WVAL-standardized N1-N2 copies/mL and WVAL-standardized N1-N2 copies/copies of PMMoV. For example, laboratory θ is generally distant from the benchmark in most units but appears among the top 5 closest laboratories when measurements are expressed as N1-N2 copies/copies of PMMoV. WVAL-standardization further changes the geometry of the PCA space, with observable changes in laboratory-benchmark distances for both N1-N2 copies/mL and N1-N2 copies/copies of PMMoV. In some cases, laboratories appear relatively close to the benchmark before standardization and shift farther after WVAL adjustment, while others show more modest changes. Despite these shifts, some laboratories show relatively stable patterns, with laboratories τ and ψ generally positioned closer to the benchmark, and laboratories η and ξ more distant across most units.

**Figure 3:**
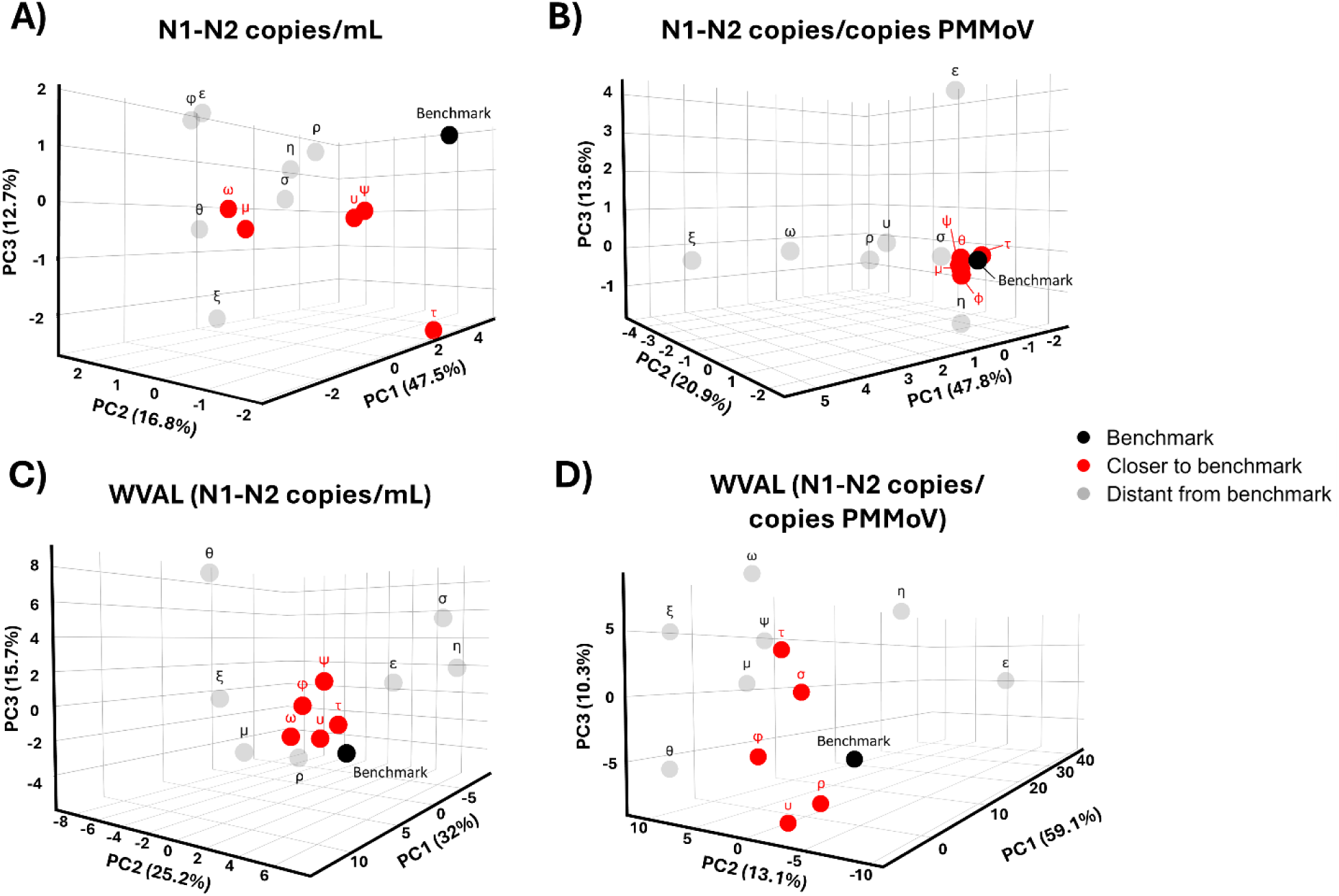
Principal Component Analysis (PCA) score plots of inter-laboratory wastewater SARS-CoV-2 measurements across the four WEM units. Each panel displays laboratory-level summarized measurements projected onto the first two PCs (PC1 and PC2) for: A, B) N1-N2 copies/mL and N1-N2 copies/copies of PMMoV; C, D) WVAL-standardized N1-N2 copies/mL; and WVAL-standardized N1-N2 copies/copies of PMMoV. PC1 and PC2 capture the dominant axes of variation among laboratories, and the relative distance between laboratories and the benchmark reflects differences in their multivariate measurement patterns.

Table 1 reports PCA-based Euclidean distances computed using the first three PCs, between each laboratory and the benchmark across the four measurement units, with the five closest laboratories highlighted in each case. These rankings are intended as descriptive summaries, as standard PCA does not provide uncertainty estimates for PC scores or distances. Across units, laboratories τ, υ, φ, and ψ appear among the five closest to the benchmark in at least three of the four cases, indicating relatively consistent proximity across measurement definitions. In contrast, laboratories ρ, θ, σ, ω, and μ exhibit more unit-specific variation, while laboratories ε, η, and ξ are not ranked among the five closest under any unit and generally remain farther from the benchmark.

**Table 1:** Summary of interlaboratory performance across three complementary concordance measures with the benchmark WEM measurement for N1-N2 copies/mL, N1-N2 copies/copies of PMMoV, WVAL-standardized N1-N2 copies/mL and WVAL-standardized N1-N2 copies/copies of PMMoV. Cohen’s к quantifies agreement in viral measurement activity classification relative to the benchmark, Euclidean distance summarizes laboratory proximity within principal component space derived from PCA, and the Exponentiated Deviation reflects relative effect size estimates from the mixed-effects model. Highlighted grey cells indicate laboratories categorized as “good” performing laboratories, orange cells indicate “poor” performing laboratories, while unhighlighted cells are categorized as “moderate” performing laboratories.

| N1-N2 copies/mL |  |  |  |
| --- | --- | --- | --- |
| Method | Cohen's $\kappa$ | Euclidean Distance | Exponentiated Deviation |
| $\varepsilon$ | 0.42<sup>α</sup> | 8.32<sup>β</sup> | -79.3%<sup>$\gamma$</sup> |
| $\eta$ | 0.93 | 8.00<sup>β</sup> | -74.4%<sup>$\gamma$</sup> |
| $\theta$ | 0.24<sup>α</sup> | 9.21<sup>β</sup> | -82.2%<sup>$\gamma$</sup> |
| $\mu$ | 0.69 | 6.82 | -45.9%<sup>$\gamma$</sup> |
| $\xi$ | 0.39<sup>α</sup> | 7.81<sup>β</sup> | -57.2%<sup>$\gamma$</sup> |
| $\rho$ | 0.76 | 7.38<sup>β</sup> | -77.0%<sup>$\gamma$</sup> |
| $\sigma$ | 0.80 | 6.92<sup>β</sup> | -63.6%<sup>$\gamma$</sup> |
| $\tau$ | 0.75 | 5.71 | -30.0% |
| $\upsilon$ | 0.73 | 6.62 | -55.5%<sup>$\gamma$</sup> |
| $\varphi$ | 0.46<sup>α</sup> | 7.59<sup>β</sup> | -55.6%<sup>$\gamma$</sup> |
| $\psi$ | 0.79 | 3.14 | 1.7% |
| $\omega$ | 0.51<sup>α</sup> | 6.42 | -37% |

| N1-N2 copies/copies of PMMoV |  |  |  |
| --- | --- | --- | --- |
| Method | Cohen's $\kappa$ | Euclidean Distance | Exponentiated Deviation |
| $\varepsilon$ | 0.40<sup>α</sup> | 5.42<sup>β</sup> | 277.1%<sup>$\gamma$</sup> |
| $\eta$ | 1.00 | 2.26<sup>β</sup> | 186.7%<sup>$\gamma$</sup> |
| $\theta$ | 0.24<sup>α</sup> | 0.75 | 5.9% |
| $\mu$ | 0.46<sup>α</sup> | 0.51 | 81.2% |
| $\xi$ | 0.13<sup>α</sup> | 7.80<sup>β</sup> | 627.1%<sup>$\gamma$</sup> |
| $\rho$ | 0.58 | 5.48<sup>β</sup> | 384.8%<sup>$\gamma$</sup> |
| $\sigma$ | 0.62 | 1.69<sup>β</sup> | 193.7%<sup>$\gamma$</sup> |
| $\tau$ | 0.63 | 0.56 | -34.4% |
| $\upsilon$ | 0.41<sup>α</sup> | 2.86<sup>β</sup> | 363.3%<sup>$\gamma$</sup> |
| $\varphi$ | 0.44<sup>α</sup> | 0.639 | 80.9% |
| $\psi$ | 0.77 | 0.49 | 90.4% |
| $\omega$ | 0.59 | 7.62<sup>β</sup> | 1088.1%<sup>$\gamma$</sup> |

| WVAL-Standardized N1-N2 copies/mL |  |  |  |
| --- | --- | --- | --- |
| Method | Cohen's $\kappa$ | Euclidean Distance | Exponentiated Deviation |
| $\varepsilon$ | 0.35<sup>α</sup> | 7.97<sup>β</sup> | 18.7% |
| $\eta$ | 0.84 | 9.51<sup>β</sup> | 67.7%<sup>$\gamma$</sup> |
| $\theta$ | 0.39<sup>α</sup> | 16.4<sup>β</sup> | 51%<sup>$\gamma$</sup> |
| $\mu$ | 0.67 | 15.8<sup>β</sup> | 50.1% |
| $\xi$ | 0.37<sup>α</sup> | 10.1<sup>β</sup> | 20.9% |
| $\rho$ | 0.86 | 7.25<sup>β</sup> | -12.5% |
| $\sigma$ | 0.63 | 11.6<sup>β</sup> | 51.2%<sup>$\gamma$</sup> |
| $\tau$ | 0.83 | 6.42 | 40.2% |
| $\upsilon$ | 0.69 | 2.89 | 2.1% |
| $\varphi$ | 0.60 | 4.53 | 67.3%<sup>$\gamma$</sup> |
| $\psi$ | 0.69 | 6.33 | 46.8% |
| $\omega$ | 0.58 | 4.77 | 40.7% |

| WVAL-Standardized N1-N2 copies/copies of PMMoV |  |  |  |
| --- | --- | --- | --- |
| Method | Cohen's $\kappa$ | Euclidean Distance | Exponentiated Deviation |
| $\varepsilon$ | 0.31<sup>α</sup> | 47.8<sup>β</sup> | 58.8% |
| $\eta$ | 0.93 | 9.77<sup>β</sup> | 96.5%<sup>$\gamma$</sup> |
| $\theta$ | 0.28<sup>α</sup> | 18.1<sup>β</sup> | 73.3%<sup>$\gamma$</sup> |
| $\mu$ | 0.55 | 10.7<sup>β</sup> | 48.7% |
| $\xi$ | 0.30<sup>α</sup> | 20.8<sup>β</sup> | 108%<sup>$\gamma$</sup> |
| $\rho$ | 0.66 | 4.84 | 23.6% |
| $\sigma$ | 0.58 | 5.63 | 87.3%<sup>$\gamma$</sup> |
| $\tau$ | 0.67 | 9.14 | 105.4%<sup>$\gamma$</sup> |
| $\upsilon$ | 0.55<sup>α</sup> | 7.95 | -13.1% |
| $\phi$ | 0.50 | 9.36 | 47.1% |
| $\psi$ | 0.65 | 11.6<sup>β</sup> | 103.2%<sup>$\gamma$</sup> |
| $\omega$ | 0.51 | 14.5<sup>β</sup> | 147.6%<sup>$\gamma$</sup> |

*Note: $\alpha$ : Cohen's $\kappa \leq 0.500$ ; $\beta$ : Distant Euclidean distance from the benchmark; $\gamma$ : Significant exponentiated deviation from the benchmark ( $p < 0.05$ )*

Overall, these results suggest that inter-laboratory distances in the PCA space depend on the measurement unit, with a subset of laboratories showing more consistent patterns across units. Two-dimensional views based on the first three PCs are provided in the Supplementary Material (Figure S3 in Supplementary Material).

### 3.4 Quantification of Laboratory Contributions to Measurement Variability with Mixed Effects Model

To determine whether the participating laboratories influence the measured SARS-CoV-2 measurements after accounting for the interlaboratory study design, including the hierarchical structure of biological replicates and technical replicates, a mixed-effects modelling was conducted across all four viral measurement units (N1-N2 copies/mL, N1-N2 copies/copies of PMMoV, WVAL-standardized N1-N2 copies/mL, and WVAL-standardized N1-N2 copies/copies of PMMoV). The corresponding estimated coefficients for the 12 participating laboratories across all four measurement units ranged between -1.726 and 2.475 (Table S4 in Supplementary Material).

Across all four models corresponding to each respective measurement metric, the mixed-effect analyses provide consistent evidence that the distinct laboratory methods significantly influence the interlaboratory SARS-CoV-2 WEM measurements from the source facility. In each model, inclusion of method as a fixed effect significantly improved model fit relative to models containing only the intercept and random effects, indicating a significant relationship between method and log-transformed measurements. For both the non-normalized N1-N2 copies/mL and PMMoV-normalized N1-N2 copies/copies of PMMoV, the difference between the laboratory methods was highly statistically significant (F-test, *p* < 0.0001). Similarly, models using WVAL-standardized N1-N2 copies/mL and WVAL-standardized N1-N2 copies/copies of PMMoV displayed statistically significant effects of methodology (F-test, *p* < 0.0002 and *p* = 0.0322, respectively). It is noteworthy that both WVAL-standardized measurement units displayed lower significance compared to N1-N2 copies/mL and N1-N2 copies/copies of PMMoV, suggesting the ability of WVAL standardization to moderately adjust for differences between laboratory methods; however, it does not eliminate the interlaboratory differences in measured SARS-CoV-2 values. Thus, the results provide evidence against the null hypothesis that including methods as a predictor in the model does not improve model fit to the data.

The estimated regression coefficients and their corresponding confidence intervals demonstrate that deviations from the benchmark vary both across the 12 laboratory methods and across the four measurement units (Figure 4). By design, these coefficients estimate the expected change in the log-transformed measurements associated with changing the method from the benchmark. For example, when measuring the viral concentration in N1-N2 copies/mL (Model 1), the estimated coefficient for laboratory φ is -0.813, indicating a decrease of 0.813 in log-transformed viral measurement when changing the method used to generate the measurement from the benchmark to method φ. To interpret these values on the original scale of the metric, the exponentiated coefficient is used (e^-0.813^ = 0.444), which implies that measurements from method φ are, on average, approximately 56% (1 – 0.444 = 0.556) lower than those from the benchmark for this respective metric, while holding all other factors constant. The associated *p*-value is a test of the null hypothesis that the true value of the coefficient is zero, i.e. that there is no difference in the measured viral load between the method and the benchmark. All laboratory methods have at least one measurement unit for which there is at least moderate evidence (*p* < 0.05) against the null hypothesis of no difference between the laboratory and the benchmark (Table 1). Specifically, there are three methods that are only significantly different from the benchmark in one of the four units. Those include method τ (in WVAL-standardized N1-N2 copies/copies of PMMoV); method ψ (in WVAL-standardized N1-N2 copies/copies of PMMoV), and method μ (in N1-N2 copies/copies of PMMoV) (Table 1). Furthermore, method ψ further shows weak evidence of a significant difference from the benchmark (*p*-value between 0.05 and 0.10) with N1-N2 copies/copies of PMMoV and WVAL-standardized copies/mL (Table S4 in Supplementary Material). Thus, according to this analysis, it can be concluded that methods τ, μ and ψ perform closest to the benchmark (though not for all four units). Conversely, the coefficients associated with laboratories η and σ have associated p-values of under 0.05 across all four models, indicating moderate evidence against the null hypothesis of no difference between these labs and the baseline.

**Figure 4:**
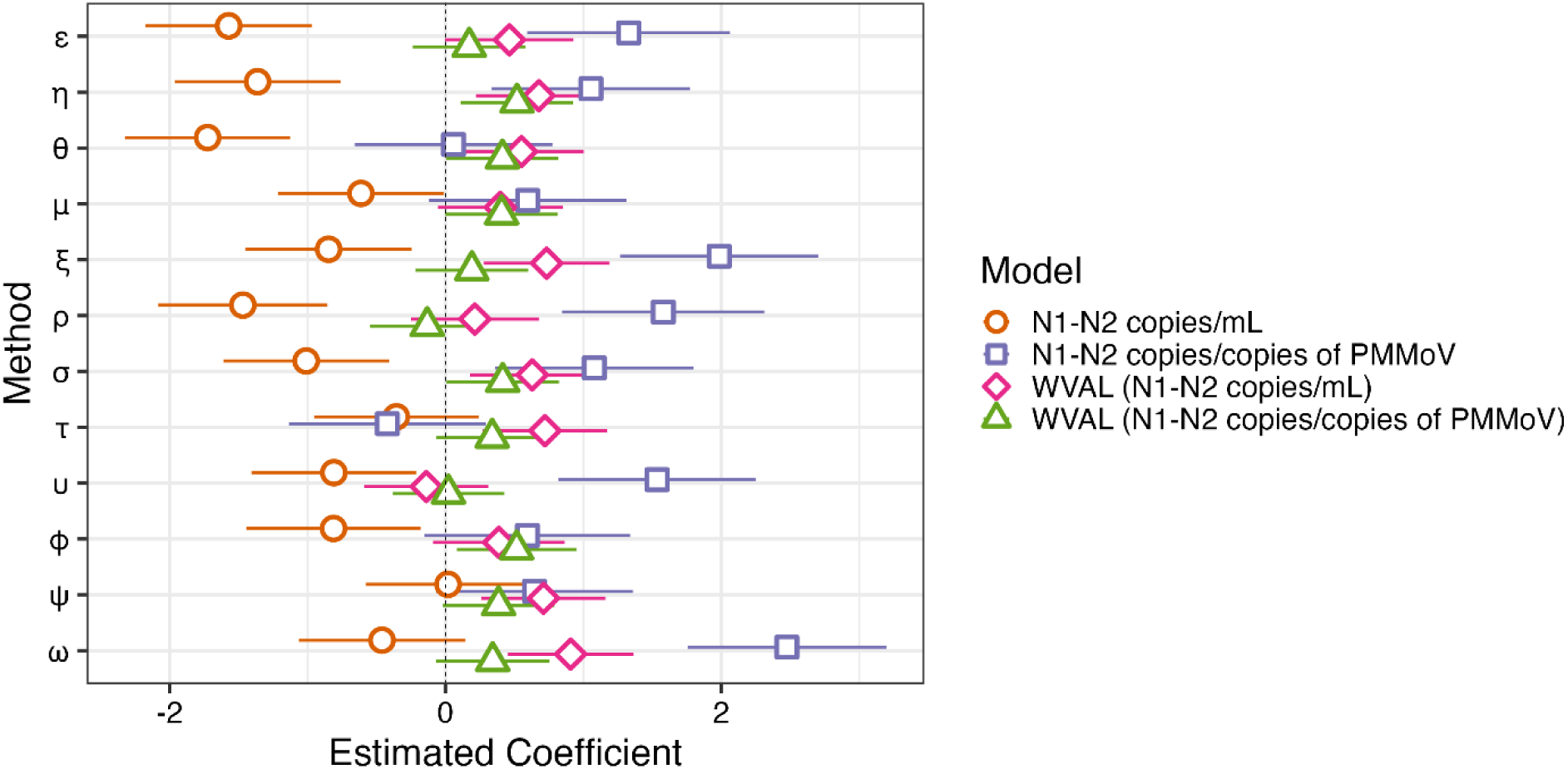
Estimated regression coefficients and corresponding 95% confidence intervals of inter-laboratory wastewater SARS-CoV-2 measurements across the four measurement units: N1-N2 copies/mL, N1-N2 copies/copies of PMMoV, WVAL-standardized N1-N2 copies/mL, and WVAL-standardized N1-N2 copies/copies of PMMoV. For each metric, the natural logarithm transform of the viral measurement was modelled. Each estimated coefficient represents the expected difference in log viral measurement for the respective laboratory compared to the benchmark, holding all other variables constant.

### 3.5 Methodological Drivers

To assess whether methodological factors contributed to interlaboratory differences in concordance with the benchmark measurement activity levels, laboratory-level associations were examined between each laboratory’s categorical concordance (match vs non-match relative to the benchmark) and the following methodological characteristics: concentration method (filtration or centrifugation), centrifugation speed (if applicable) for both spins, standardized materials in assays for the N1 and N2 gene regions, N1 and N2 RNA template volume (μL), N1 and N2 total RT-qPCR reaction volume (μL), N1 and N2 RT-step at 25°C, cDNA synthesis duration (min), initial denaturation duration (min), cycling number, cycling denaturation duration (s), and cycling annealing extension (°C), using Fisher’s exact test (as explained in Section 2.3.4). In this study, the laboratory-level comparisons did not identify a significant association between benchmark concordance and any of the methodological variables analyzed, including extracted mass (Table S2 in Supplementary Material). While no statistically significant associations were detected, the following variables pertaining to reaction setup and thermal cycling exhibited comparatively lower p-values and may merit further investigation in studies specifically designed to assess methodological drivers: RNA template volume (*p* = 0.127), total RT-qPCR reaction volume (*p* = 0.182), extracted mass at threshold between 58 to 68 mg (*p* = 0.250), and standardized assay materials for N1 and N2 gene targets (*p* = 0.250) (Table S5 in Supplementary Material). Previous RT-qPCR optimization studies have demonstrated that increasing RNA template volume can help improve the sensitivity of measurement quantification ^64^; there is currently no literature that indicates the extent of RT-qPCR reaction volume on SARS-CoV-2 N1 and N2 quantification. For the standard material, in this study, 11 of the 12 participating laboratories used the RNA Exact Diagnostics (EDX) SARS-CoV-2 standard (Bio-Rad Laboratories), a commercially produced synthetic RNA transcript (Table S2 in the Supplementary Material). In contrast, only 1 laboratory (method θ) used its in-house concatenated plasmid, a laboratory-constructed plasmid DNA standard containing concatenated SARS-CoV-2 N1 and N2 gene target regions (Table S2 in the Supplementary Material). There is no literature directly comparing them as quantification standards for N1 and N2 SARS-CoV-2 quantification. However, by analogy with other RNA versus laboratory-constructed plasmid standards, EDX and in-house concatenated plasmid are expected to yield different absolute SARS-CoV-2 copy measurements through their impact on standard curves (e.g., differences in molecular form, inclusion or omission of the reverse transcription step, and associated amplification efficiency effects) ^32,65^. For the RT-qPCR master mix, cDNA synthesis duration, and initial denaturation duration, there is general evidence that these parameters can influence sensitivity and assay efficiency, but their impacts are typically mitigated when using commercial kits within the manufacturer’s guidelines ^64^.

Among the methodological variables evaluated, extracted mass was the only parameter estimated at the individual sample level rather than solely at the laboratory level (Table S3 in Supplementary Material). This provided an opportunity for further assessment of its potential on benchmark concordance beyond the laboratory-level analysis described above. Furthermore, because SARS-CoV-2 RNA is known to preferentially partition to wastewater solids, often exhibiting orders-of-magnitude higher concentrations in primary settled solids than in the liquid fraction, extracted mass is a plausible driver of interlaboratory variability for SARS-CoV-2 measurements ^66,67^. Extracted mass was therefore further evaluated at the sample level (interlaboratory Samples A to M), examining whether dichotomizing mass across a range of thresholds was associated with benchmark concordance.

For the N1-N2 copies/mL and N1-N2 copies/copies of PMMoV, p-values remained above 0.05, ranging between 0.230 to 1.000 at extracted mass thresholds ranging between 25 to 250 mg with high sensitivity to threshold changes (Figure S4 in Supplementary Material). However, it is noteworthy some thresholds exhibited comparatively lower p-values, specifically at extracted mass thresholds of 47 and 49 mg in N1-N2 copies/mL, and 25, 26, and 160 mg (*p* < 0.250) (Figure S4 in Supplementary Material). There are no studies to date explicitly elucidating the impact of extracted mass input for RNA extraction to N1 and N2 quantification quality or sensitivity. This study was limited to observational analyses of the interlaboratory data that was not specifically designed to isolate extracted mass effects on benchmark concordance. Future work should employ controlled experiments varying pellet weight input to assess impacts on N1 and N2 measurement precision.

### 3.6 Summary of Results

Across all three analytical approaches, a consistent pattern emerges: laboratory methods meaningfully influence both SARS-CoV-2 viral concentration measurements and their downstream interpretation in public health. The WEM viral activity classification comparison between methods and benchmark heatmaps reveals where disagreements arise, particularly during periods of fluctuating wastewater measurement, while demonstrating strong convergence at epidemiological extremes (very low or very high activity) (Figure 2). PCA further demonstrates that laboratories differ not only in magnitude but in multivariate measurement profiles, with some methods clustering tightly around the benchmark and others showing systematic divergence across measurement units (Figure 3). Mixed-effects modelling confirms that laboratory identity is a statistically significant predictor of viral measurement across all four units, even after accounting for replicate structure, indicating that methodological choices potentially affect quantitative outputs (Figure 4). Taken together, these analyses demonstrate that methodological variability propagates into interpretive variability, underscoring the need to identify specific analytical drivers of divergence and to advance harmonization strategies within WEM programs.

For N1-N2 copies/mL, laboratory methods exhibited higher variability from the benchmark across all three analytical concordance measures (Table 1). A subset of methods, in particular methods τ, υ, and ψ demonstrated consistently strong agreement, with high Cohen’s к (к > 0.5) and small Euclidean distance or no significant exponentiated deviation and were therefore classified as good-performing methods. In contrast, several methods, particularly methods ε, θ, ξ, and φ, exhibited poor agreement with the benchmark across all three analytical methods, characterized by low Cohen’s к (≤ 0.5), large Euclidean distance, and significant exponentiated deviations, and were classified as poor-performing laboratories. The remaining methods (methods η, μ, ρ, σ, υ, and ω) displayed mixed performance, typically demonstrating acceptable agreement by one or two analytical methods, but pronounced deviation by others, and were classified as moderate performing laboratories. Overall, the N1-N2 copies/mL measurement displayed the greatest spread in laboratory agreement relative to the benchmark. For N1-N2 copies/copies of PMMoV, performance measures became slightly attenuated, but interlaboratory divergence persisted (Table 1). In this case, laboratories τ and ψ demonstrated consistently strong concordance to the benchmark across all three analytical methods and were therefore classified as high-performing laboratories. In contrast, laboratories ε, ξ, and υ demonstrated consistently poor concordance with the benchmark across all three analytical methods (Table 1). The remaining laboratories (laboratories η, θ, μ, ξ, ρ, σ, φ, and ω) displayed mixed performance and were classified as moderate performing laboratories (Table 1). These results indicate that PMMoV normalization improved the concordance of some laboratories to the benchmark (laboratories θ, ξ, and φ) while amplifying the divergence for laboratory υ (Table 1).

For WVAL-standardized N1-N2 copies/mL, concordance with the benchmark improved for various laboratories that previously displayed poor concordance with the benchmark with the non-WVAL-standardized N1-N2 copies/mL (laboratories ξ and φ). Laboratories τ, υ, and ψ demonstrated consistently strong concordance to the benchmark, classifying them as high-performing laboratories for this measurement metric (Table 1). Laboratory θ was the only laboratory that exhibited low Cohen’s к, high Euclidean distance, and significant exponentiated deviation from the benchmark, making it the only poor-performing laboratory for this measurement metric (Table 1), while the remaining laboratories (laboratories ε, η, μ, ξ, ρ, σ, and φ) fell into the moderate-performing category showing mixed performance across all three analytical concordance measures (Table 1). Compared to the non-WVAL standardized N1-N2 copies/mL, WVAL-standardization reduced the number of laboratories exhibiting significant deviation across all three analytical methods. Lastly, for WVAL-standardized N1-N2 copies/copies of PMMoV, although Cohen’s к generally indicated moderate to strong agreement to the benchmark (≥ 0.5) for 10 out of the 12 participating laboratories, large Euclidean distances were exhibited for 8 out of the 12 laboratories, and significant exponentiated deviation for 7 out of the 8 laboratories (Table 1), resulting in relatively few laboratories meeting criteria for good performance across all three analytical methods, with only laboratories ρ and φ being categorized as high performing laboratories. Laboratories θ and ξ exhibited consistently lower performance relative to the benchmark for all concordance measures, classifying them as poor-performing laboratories, while the remaining laboratories (laboratories ε, η, μ, σ, τ, υ, ψ, and ω) exhibited mixed performance across all three analytical methods, classifying them as moderate laboratories (Table 1).

These discrepancies in laboratory classification across the three analytical methods are likely driven by differences in data processing of the interlaboratory data in this study. Firstly, the approach applying Cohen’s к relies on categorizing a continuous viral measurement into discrete agreement classes (i.e., low, moderate, high, and very high), making them inherently sensitive to the choice of cut-off values used to define benchmark concordance. In contrast, the PCA analysis operates on centered and scaled continuous data for both N1-N2 copies/mL and N1-N2 copies/copies of PMMoV, emphasizing relative differences from the benchmark rather than measurement categorization. Lastly, the mixed effects model approach applies a natural logarithmic transformation to viral measurements to satisfy model assumptions. The effect of this logarithmic transformation is non-uniform, compressing differences among larger values while preserving relative differences among smaller values. For example, reducing a difference between values of 10 and 20 from 10 units to 0.60 after logarithmic transformation, while a difference between values of 0.8 and 1.1 remains 0.3 with and without logarithmic transformation. Finally, methodological differences in data aggregation further contribute to divergence across the three analytical approaches. The approach applying Cohen’s к and the PCA analysis both rely on averaged biological replicates for each sample reported in the interlaboratory analysis, whereas the mixed-effects model retains replicate-level measurements and explicitly accounts for additional variability through random effects.

## 4 Limitations

This study has limitations that should be considered when interpreting its findings. Firstly, the interlaboratory data throughout this study were observational rather than experimentally controlled. Participating laboratories applied their routine SARS-CoV-2 methods without harmonized methodological constraints, and metadata availability varied across participating laboratory methods and across interlaboratory samples. As a result, attribution of interpretive differences to specific methodological steps was not possible. The limited granularity of the methodological metadata, particularly regarding wastewater solids concentration, nucleic acid extraction, and RT-qPCR parameters, restricted the ability to isolate the specific contributions of the individual analytical steps beyond extracted solids mass.

Secondly, although the benchmark derived from the source facility’s longitudinal WEM dataset was able to be validated against clinical case data within the exact sewershed of the collected wastewaters, it provides a practical and epidemiologically grounded reference point, and it does not represent a true concentration. Endogenous wastewater samples in this study lacked a definitive “true” concentration, and the benchmark reflected expected measurement patterns specific to the sampling site. Thus, concordance with the benchmark should be interpreted as convergence toward a locally consistent measurement rather than accuracy relative to an absolute standard. Interlaboratory studies that attempt to establish a “true” concentration through spiked (externally added) virus or surrogate are likewise challenging, because spike-and-recovery experiments may not adequately represent the behaviour or recovery of authentic endogenous SARS-CoV-2 in heterogeneous wastewater matrices ^68^.

Finally, the chi-square test that was performed to assess methodological drivers (Section 3.6) assumes that observations are independent, which is unlikely given that repeated measurements use the same method. Addressing this limitation would require data from a well-designed experiment rather than purely observational data or further modelling approaches that explicitly account for interlab dependence.

## 5 Conclusions and Recommendations

This study demonstrates that differences between 12 different laboratory methods used across a provincial WEM network can lead to substantially different measurement magnitudes and potential misclassification of viral activity levels for public health interpretation from the same source facility. Using an endogenous interlaboratory dataset that was processed under routine operational workflows of the Ontario WSI program, this study demonstrates that method-specific approaches lead the divergence in SARS-CoV-2 concentrations relative to a facility-specific longitudinal benchmark. Across the three concordance metrics approaches, laboratory method was a statistically significant driver of differences in reported WEM measurements. Stronger concordance between the laboratory methods and the benchmark was exhibited during epidemiological extremes of the COVID-19 waves (very low or very high wastewater measurement), but substantial interpretive divergence occurred during the transitional periods, when WEM remain informative for guiding public health responses. While this study identified extracted wastewater mass as a potential driver for concordance between participating laboratories and the benchmark, this study was limited by the observational nature of the interlaboratory data, requiring controlled experimental work to further isolate and quantify the contribution of the individual methodological components.

This study provides evidence that laboratory methods used in routine SARS-CoV-2 WEM can materially shape public health interpretations. As WEM continues to expand beyond SARS-CoV-2 into multi-pathogen monitoring, addressing interpretive variability is essential to ensure reliable, comparable, and actionable surveillance data.

## Acknowledgements

The authors would like to acknowledge the Ontario Ministry of the Environment, Conservation and Parks (MECP). The work was made possible by the ongoing support from the Ontario Wastewater Surveillance Consortium (OWSC) and the member institutions across Ontario, Canada, which was established under the leadership of MECP. The OWSC played a pivotal role in the data acquisition, quality assurance, and methodological development for all the interlaboratory data used in this study. The authors gratefully acknowledge the vital contributions of all employees from the Province of Ontario, the MECP, and the Ontario Ministry of Health. This work was also supported by the CIHR Applied Public Health Chair in Environment, Climate Change and One Health, awarded to Robert Delatolla, the Natural Sciences and Engineering Research Council of Canada (NSERC) Vanier Canada Graduate Scholarship awarded to Nada Hegazy, and the Canadian Statistical Sciences Institute (CANSSI).

## Data Availability

The datasets generated and analyzed in this study are not publicly available due to data governance restrictions but are available from the authors on request.

