## Supplementary material for "Twelve Distinct Laboratory Methods Used to Measure SARS-CoV-2 in Wastewaters throughout a Three-Year Ontario-Wide, Canada Study: Impact on Public Health Interpretation of Disease Incidence": Supplmentary Material

**Table S1:** Summary of laboratory-specific solids concentration methods used in the interlaboratory study by the participating laboratories during the study period.

| Laboratory | Extracted mass (mg) | Concentration method | Centrifugation conditions (at 4°C) | Extraction Kit |
|---|---|---|---|---|
| **ε** | 28.64 | Centrifugation | Mar. 24, 2021 – Mar. 31, 2024<br>Spin 1: 27,792 x g (90 min)<br>No second spin<br><br>Mar. 31, 2023 – Jul. 31, 2024<br>Spin 1: 4,500 x g (120 min) | Mar. 24, 2021 – May 23, 2023: PowerMicrobiome<br>May 24, 2023 – Jul. 30, 2024: PowerFecal Pro |
| **η** | 28.64 | PEG | Spin 1: 13,000 x g (90 min)<br>No second spin | Mar. 24, 2021 – May 23, 2023: PowerMicrobiome<br>May 24, 2023 – Jul. 30, 2024: PowerFecal Pro |
| **θ** | 76.38 | Centrifugation | Spin 1: 10,000 x g (45 min)<br>Spin 2: 13,000 x g (1 min) | PowerMicrobiome |
| **μ** | Not applicable | Filtration | Not applicable | Mar. 24, 2021 – May 23, 2023: PowerMicrobiome<br>May 24, 2023 – Jul. 30, 2024: PowerViral |
| **ξ** | 91.65 | PEG | Spin 1: 12,000 x g (120 min)<br>No second spin | PowerMicrobiome |
| **ρ** | 76.38 | Centrifugation | Spin 1: 12,000 x g (50 min)<br>Spin 2: 12,000 x g (15 min) | PowerViral |
| **σ** | 76.38 | Centrifugation | Spin 1: 12,000 x g (60 min)<br>No second spin | PowerMicrobiome |
| **τ** | 250.00 | Centrifugation | Spin 1: 10,000 x g (45 min)<br>Spin 2: 10,000 x g (5 min) | Mar. 24, 2021 – May 23, 2023: PowerMicrobiome<br>May 24, 2023 – Jul. 30, 2024: PowerViral |
| **υ** | 124.11 | Centrifugation | Spin 1: 12,000 x g (60 min)<br>No second spin | PowerViral |
| **φ** | 109.40 | PEG | Spin 1: 8,000 x g (60 min)<br>No second spin | PowerViral |
| **ψ** | 122.20 | PEG | Spin 1: 12,000 x g (90 min)<br>Spin 2: 12,000 x g (5 min) | PowerMicrobiome |
| **ω** | 57.28 | Centrifugation | Spin 1: 4,200 x g (20 min)<br>Spin 2: 20,000 x g (2 min) | Mar. 24, 2021 – May 23, 2023: PowerMicrobiome<br>May 24, 2023 – Jul. 30, 2024: PowerViral |

**Table S2:** Summary of laboratory-specific RT-qPCR methods used in the interlaboratory study by the participating laboratories during the study period.

| Laboratory | PCR Platform | RT-qPCR master mix | Target genes | Standardized Assay Materials (N1, N2) | Template volume (µ) | Reaction volume | Cycling conditions |
|---|---|---|---|---|---|---|---|
| **ε** | QuantStudio | Mar. 24, 2021 – May 23, 2023: TaqPath One-Step<br><br>May 23, 2023 – Jul. 30th, 24: Luna Probe One-Step | N1, N2 | EDX | N1: 2.5<br>N2: 2.5 | N1: 10<br>N2: 10 | Reverse transcription at 50 °C for 5 min; initial denaturation at 95 °C for 20 s; 45 amplification cycles consisting of 95 °C for 15 s and 60 °C for 40 s. |
| **η** | QIAquant | Fast Virus One-Step Master Mix | N1, N2 | EDX | N1: 5<br>N2: 5 | N1: 20<br>N2: 20 | Reverse transcription at 50 °C for 15 min; initial denaturation at 95 °C for 2 min; 45 amplification cycles consisting of 95 °C for 3 s, 55 °C for 30 s (N1), and 60 °C for 30 s (N2). |
| **θ** | CFX | Fast Virus One-Step Master Mix | N1, N2 | Concatenated plasmid | N1: 4<br>N2: 4 | N1: 10<br>N2: 10 | Reverse transcription at 50 °C for 5 min; initial denaturation at 95 °C for 2 min; 45 amplification cycles consisting of 95 °C for 5 s and 60 °C for 30 s. |
| **µ** | MA6000 | Mar. 24, 2021 – Feb. 6th, 2023: Takyon Dry One-Step RT Probe<br><br>Feb. 3rd, 2023 – Jul. 30, 2024: Luna Probe One-Step | N1, N2 | EDX | N1: 5<br>N2: 5 | N1: 20<br>N2: 20 | Reverse transcription at 48–55 °C for 10 min; initial denaturation at 95 °C for 3 min; 45 amplification cycles consisting of 95 °C for 10 s and 60 °C for 30–55 s |
| **ξ** | CFX | Reliance RT-qPCR | N1, N2 | EDX | N1: 5<br>N2: 5 | N1: 15<br>N2: 15 | Reverse transcription at 50 °C for 10 min; initial denaturation at 95 °C for 10 min; 45 amplification cycles consisting of 95 °C for 10 s and 60 °C for 30 s. |
| **ρ** | CFX | Reliance RT-qPCR | N1, N2 | EDX | N1: 5<br>N2: 5 | N1: 10<br>N2: 10 | Reverse transcription at 50 °C for 10 min; initial denaturation at 95 °C for 10 min; |

| | | | | | | | |
|---|---|---|---|---|---|---|---|
| | | | | | | | 45 amplification cycles consisting of 95 °C for 30 s and 60 °C for 30 s. |
| **σ** | QIAquant | TaqMan Fast Virus One-Step | N1, N2, E | EDX | N1: 5<br>N2: 5 | N1: 10<br>N2: 10 | Reverse transcription at 50 °C for 10 min; initial denaturation at 95 °C for 10 min; 45 amplification cycles consisting of 95 °C for 5 s and 60 °C for 30 s. |
| **τ** | CFX | TaqMan Fast Virus One-Step | N1, N2 | EDX | N1: 3<br>N2: 3 | N1: 10<br>N2: 10 | Reverse transcription at 50 °C for 10 min; initial denaturation at 95 °C for 10 min; 45 amplification cycles consisting of 95 °C for 5 s and 60 °C for 30 s |
| **υ** | QuantStudio | Reliance RT-qPCR | N1, N2 | EDX | N1: 5<br>N2: 5 | N1: 20<br>N2: 20 | Reverse transcription at 50 °C for 15 min; initial denaturation at 95 °C for 10 min; 40 amplification cycles consisting of 95 °C for 10 s and 60 °C for 35 s. |
| **φ** | CFX | Reliance One-Step | N1, N2, E | EDX | N1: 5<br>N2: 5 | N1: 20<br>N2: 20 | Reverse transcription at 50 °C for 10 min; initial denaturation at 95 °C for 10 min; 45 amplification cycles consisting of 95 °C for 5 s and 60 °C for 30 s. |
| **ψ** | CFX | TaqPath One-Step | N1, N2, N200 | EDX | N1: 5<br>N2: 5 | N1: 20<br>N2: 20 | Reverse transcription at 50 °C for 15 min; initial denaturation at 95 °C for 2 min; 45 amplification cycles consisting of 95 °C for 3 s, 55 °C for 30 s (N1), and 60 °C for 30 s (N2) |
| **ω** | CFX | Reliance One-Step | N1, N2 | EDX | N1: 5<br>N2: 5 | N1: 20<br>N2: 20 | Reverse transcription at 50 °C for 10 min; initial denaturation at 95 °C for 10 min; 45 amplification cycles consisting of 95 °C for 10 s and 60 °C for 30 s. |

**Table S3:** Estimated extracted mass by each participating laboratory

| Laboratory | A | B | C | D | E | F | G | H | I | J | K | L | M |
|---|---|---|---|---|---|---|---|---|---|---|---|---|---|
| **ε** | 29.4 | 29.4 | 30.4 | 34.6 | 23.7 | 47.2 | 46.9 | 35.4 | 18.2 | 14.4 | 14.2 | 24.6 | 24 |
| **η** | 29.4 | 29.4 | 30.4 | 34.6 | 23.7 | 47.2 | 46.9 | 35.4 | 18.2 | 14.4 | 14.2 | 24.6 | 24 |
| **θ** | 78.4 | 78.4 | 81.2 | 92.2 | 63.2 | 125.8 | 125 | 94.3 | 48.5 | 38.4 | 37.9 | 65.6 | 64 |
| **μ** | NA | NA | NA | NA | NA | NA | NA | NA | NA | NA | NA | NA | NA |
| **ξ** | 58.8 | 58.8 | 60.9 | 69.2 | 47.4 | 94.4 | 93.7 | 70.7 | 36.4 | 28.8 | 28.4 | 49.2 | 48 |
| **ρ** | 78.4 | 78.4 | 81.2 | 92.2 | 63.2 | 125.8 | 125 | 94.3 | 48.5 | 38.4 | 37.9 | 65.6 | 64 |
| **σ** | 78.4 | 78.4 | 81.2 | 92.2 | 63.2 | 125.8 | 125 | 94.3 | 48.5 | 38.4 | 37.9 | 65.6 | 64 |
| **τ** | 250 | 250 | 250 | 250 | 250 | 250 | 250 | 250 | 250 | 250 | 250 | 250 | 250 |
| **υ** | 127.4 | 127.4 | 131.9 | 149.9 | 102.8 | 204.4 | 203 | 153.2 | 78.8 | 62.5 | 61.6 | 106.5 | 104 |
| **φ** | 91.5 | 91.5 | 80.7 | 107.6 | 90.7 | 104.8 | 80.7 | 52.2 | 38 | 32.2 | 22 | 43.1 | 53.8 |
| **ψ** | 78.4 | 78.4 | 81.2 | 92.2 | 63.2 | 125.8 | 125 | 94.3 | 48.5 | 38.4 | 37.9 | 65.6 | 64 |
| **ω** | 58.8 | 58.8 | 60.9 | 69.2 | 47.4 | 94.4 | 93.7 | 70.7 | 36.4 | 28.8 | 28.4 | 49.2 | 48 |

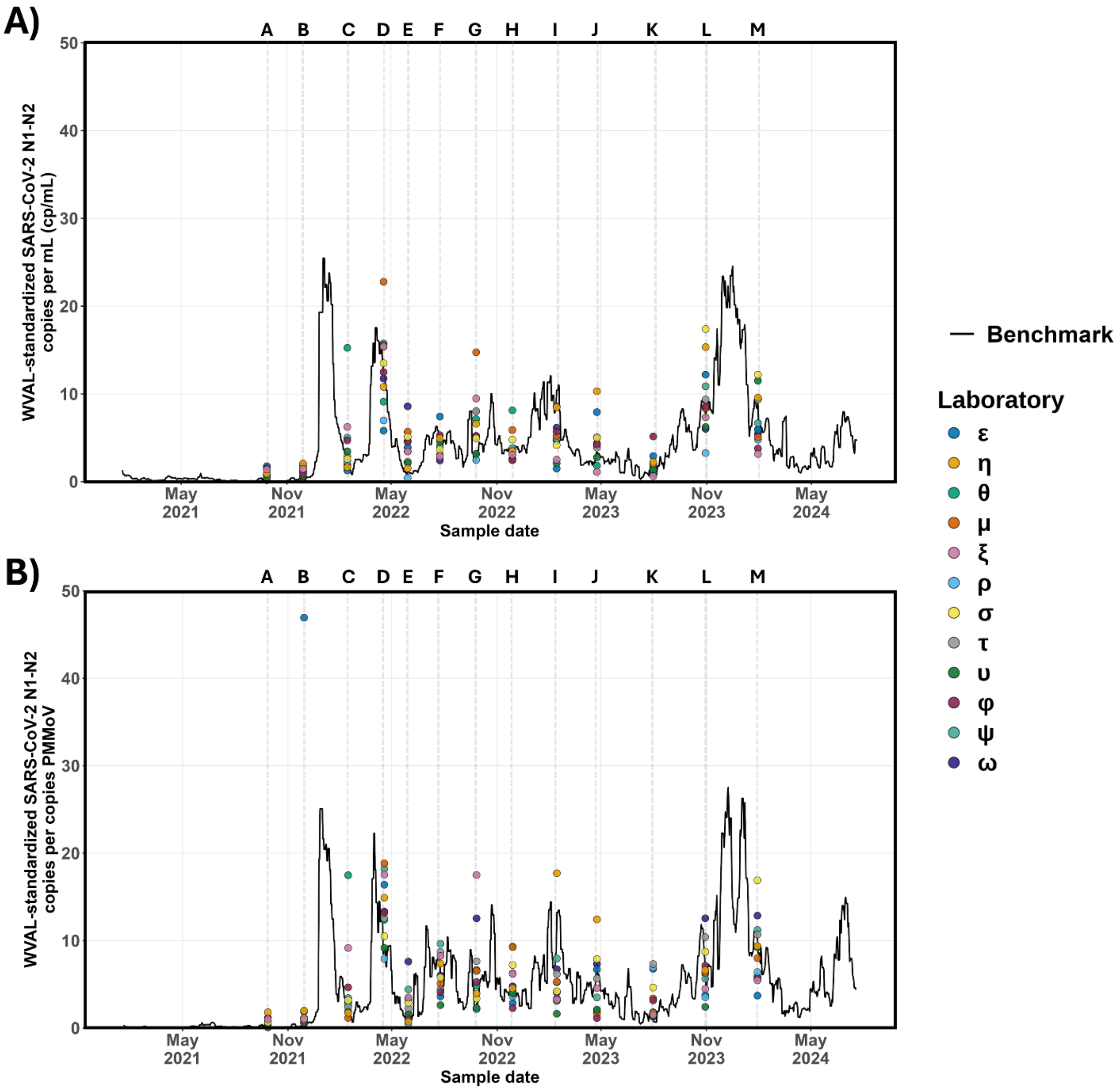


**Figure S1: Inter-laboratory measurement relative to the benchmark measurement from the source facility.** The solid black line represents the baseline measurement, defined as the 7-day moving average of A) WVAL-standardized N1-N2 copies/mL and B) WVAL-standardized N1-N2 copies/copies of PMMoV from the benchmark source facility from which the inter-laboratory samples were collected. Colored circles represent measurements reported by the 12 participating laboratories analyzing wastewater samples collected from the same location. Sample letters above the plotting area correspond to individual inter-laboratory sampling rounds.

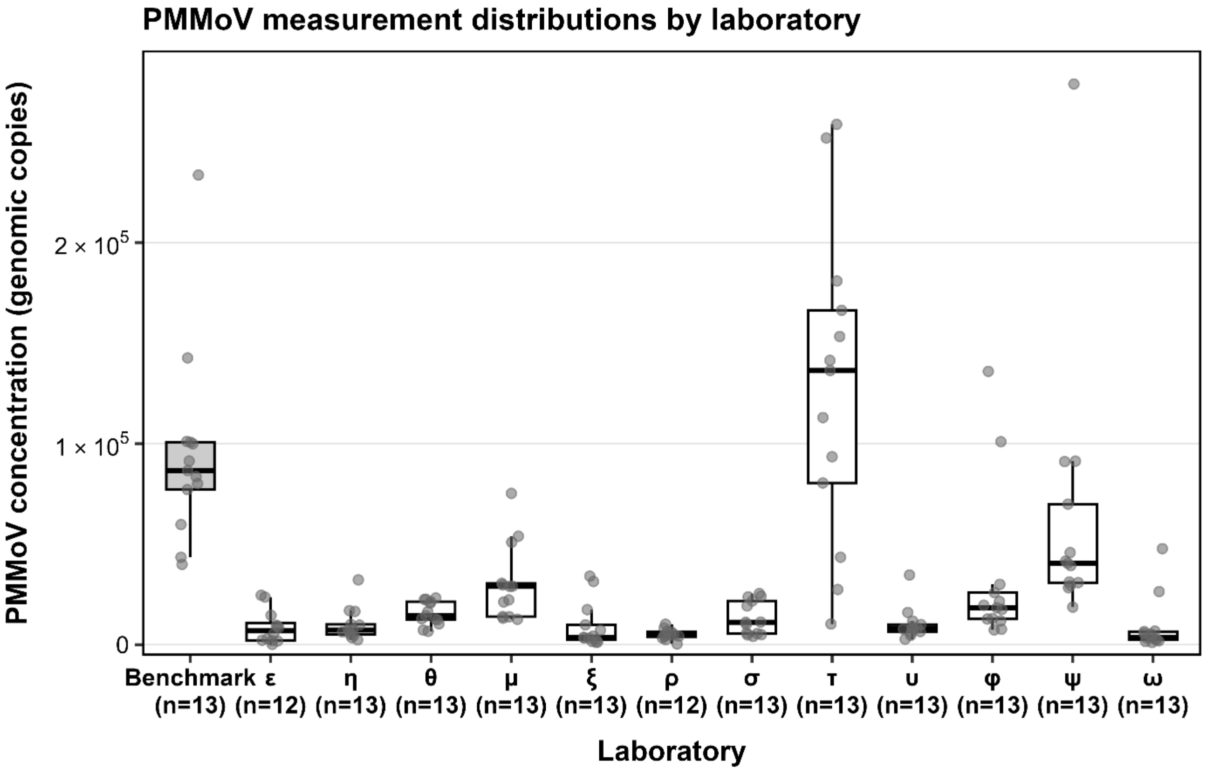


**Figure S2:** Distribution of PMMoV normalization biomarker measured by participating laboratories throughout the interlaboratory study

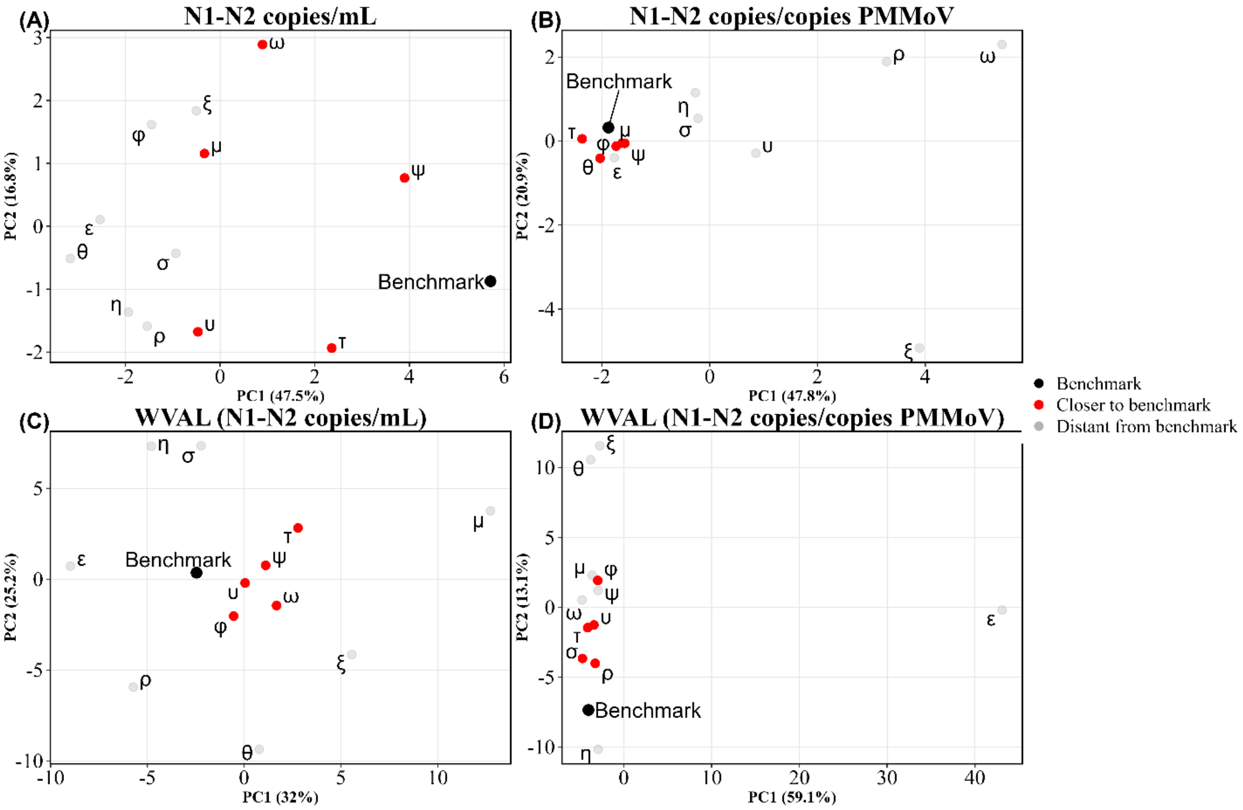


**Figure S3:** Principal Component Analysis (PCA) score plots of inter-laboratory wastewater SARS-CoV-2 measurements across the four WEM units. Each panel displays laboratory-level summarized measurements projected onto the first two PCs (PC1 and PC2) for: (A & B) N1-N2 copies/mL and N1-N2 copies per copies of PMMoV; (C & D) WVAL- standardized copies per mL; and WVAL-standardized copies per PMMoV copy. PC1 and PC2 capture the dominant axes of variation among laboratories, and the relative distance between laboratories and the benchmark reflects differences in their multivariate measurement patterns.

For laboratories 1 to 14, the estimated coefficients in Table S4 are the estimated coefficients displayed in Figure 4, which are transformed to the expected percent deviation from the baseline in Table 1. The estimated coefficients for the benchmark (intercept) were not included in Figure 4 as they are not of direct interest, since our analysis focuses on the differences between the benchmark and each laboratory. Note that the values of the intercepts vary across units due to their varying magnitudes.

**Table S4:** For each of the four models, this table includes the estimated coefficient for each laboratory (displayed in Figure 4), and the associated standard error (in parentheses), and the range of the p-value, indicated by asterisks.

| | Dependent variable | | | |
|---|---|---|---|---|
| **Laboratory** | **N1N2 (cp/mL)** | **N1N2 (cp/cp)** | **WVAL (N1N2 cp/mL)** | **WVAL (N1N2 cp/cp)** |
| **Benchmark/Intercept** | 3.818***<br>(0.371) | -7.541***<br>(0.424) | 0.901***<br>(0.267) | 0.746***<br>(0.276) |
| **ε** | -1.573***<br>(0.308) | 1.327***<br>(0.375) | 0.171<br>(0.209) | 0.462*<br>(0.238) |
| **η** | -1.364***<br>(0.307) | 1.053***<br>(0.367) | 0.517**<br>(0.209) | 0.676***<br>(0.232) |
| **θ** | -1.726***<br>(0.306) | 0.057<br>(0.366) | 0.412**<br>(0.208) | 0.550**<br>(0.231) |
| **μ** | -0.614**<br>(0.306) | 0.595<br>(0.366) | 0.406*<br>(0.208) | 0.397*<br>(0.231) |
| **ξ** | -0.848***<br>(0.307) | 1.984***<br>(0.367) | 0.190<br>(0.209) | 0.732***<br>(0.232) |
| **ρ** | -1.470***<br>(0.313) | 1.578***<br>(0.374) | -0.133<br>(0.213) | 0.212<br>(0.236) |
| **σ** | -1.011***<br>(0.306) | 1.078***<br>(0.366) | 0.414**<br>(0.208) | 0.628***<br>(0.231) |
| **τ** | -0.357<br>(0.304) | -0.422<br>(0.364) | 0.338<br>(0.207) | 0.720***<br>(0.230) |
| **υ** | -0.810***<br>(0.305) | 1.533***<br>(0.364) | 0.021<br>(0.207) | -0.140<br>(0.230) |
| **φ** | -0.813**<br>(0.322) | 0.592<br>(0.381) | 0.515**<br>(0.221) | 0.386<br>(0.244) |
| **ψ** | 0.017<br>(0.304) | 0.644*<br>(0.364) | 0.284*<br>(0.206) | 0.709***<br>(0.230) |
| **ω** | -0.462<br>(0.307) | 2.475***<br>(0.367) | 0.341<br>(0.209) | 0.907***<br>(0.232) |
| | *Note:* $^{*}p < 0.1$; $^{**}p < 0.05$; $^{***}p < 0.01$ | | | |

**Table S5:** Exploratory associations between methodological parameters and deviation from benchmark measurement

| **Methodological Parameter** | ***p*-value** |
|---|---|
| Filtration | 1.00 |
| Centrifugation (spin 1) | 1.00 |
| Centrifugation (spin 2) | 1.00 |
| Standardized materials in assays (N1 and N2 gene regions) | 0.25 |
| Targeted gene regions | 1.00 |

| | |
|---|---|
| N1 RNA template volume (µL) | 0.127 |
| N2 RNA template volume (µL) | 0.127 |
| N1 Total RT-qPCR reaction volume (µL) | 0.182 |
| N2 Total RT-qPCR reaction volume (µL) | 0.182 |
| N1 and N2 RT-step at 25℃ | 0.510 |
| cDNA synthesis duration (min) | 0.509 |
| Initial denaturation duration (min) | 0.523 |
| Cycling number | 1.00 |
| Cycling denaturation duration (s) | 1.00 |
| Cycling annealing extension (℃) | 1.00 |
| Extracted mass at 58 – 68 mg threshold (mg) | 0.236 |

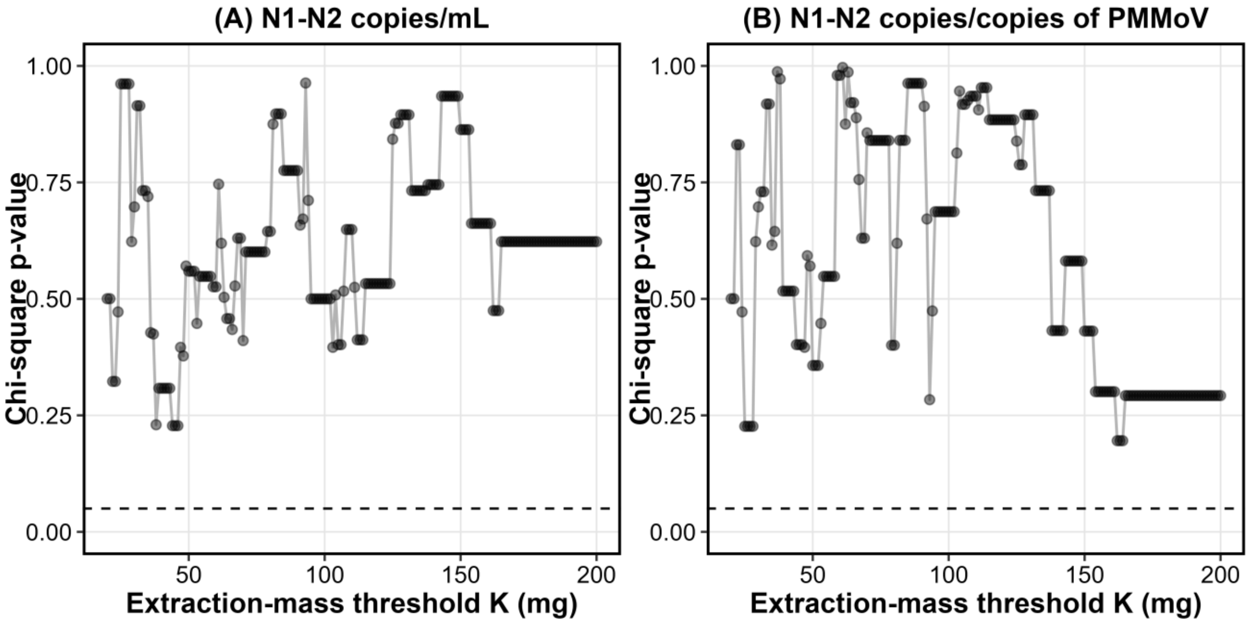


**Figure S4:** P-values from chi-square tests assessing the association between extracted mass and benchmark concordance across extraction-mass thresholds K. The x-axis shows the threshold K (mg) used to dichotomize extracted mass, and the y-axis shows the corresponding p-values. Each point represents one test; the connecting line is included only to aid visual inspection. A) N1–N2 copies/mL, and B) N1–N2 copies/copies PMMoV. The dashed horizontal line indicates the $p = 0.05$ significance level.
